# Plant to animal protein ratio in the diet: nutrient adequacy, long-term health and environmental pressure

**DOI:** 10.1101/2022.05.20.22275349

**Authors:** Hélène Fouillet, Alison Dussiot, Elie Perraud, Juhui Wang, Jean-François Huneau, Emmanuelle Kesse-Guyot, François Mariotti

## Abstract

**Background:** Animal and plant protein sources have contrasting relationships with nutrient adequacy and long-term health, and their adequate ratio is highly debated.

**Objective:** We aimed to explore how the percentage of plant protein in the diet (%PP) relates to nutrient adequacy and long-term health but also to environmental pressures, to determine the adequate and potentially optimal %PP values.

**Methods:** Observed diets were extracted from the dietary intakes of French adults (INCA3, n=1 125). Using reference values for nutrients and disease burden risks for foods, we modeled diets with graded %PP values that simultaneously ensure nutrient adequacy, minimize long-term health risks and preserve at best dietary habits. This multi-criteria diet optimization was conducted in a hierarchical manner, giving priority to long-term health over diet proximity, under the constraints of ensuring nutrient adequacy and food cultural acceptability. We explored the tensions between objectives and identified the most critical nutrients and influential constraints by sensitivity analysis. Finally, environmental pressures related to the modeled diets were estimated using the AGRIBALYSE database.

**Results:** We find that nutrient-adequate diets must fall within the ∼15%-80% %PP range, a slightly wider range being nevertheless identifiable by waiving the food acceptability constraints. Fully healthy diets, also achieving the minimum-risk exposure levels for both unhealthy and healthy foods, must fall within the 25%-70% %PP range. All of these healthy diets were very distant from current typical diet. Those with higher %PP had lower environmental impacts, notably on climate change and land use, while being as far from current diet.

**Conclusions:** There is no single optimal %PP value when considering only nutrition and health, but high %PP diets are more sustainable. For %PP>80%, nutrient fortification/supplementation and/or new foods are required.

## Introduction

Historical and current nutritional transitions are coupled with changes in the relative contribution of dietary animal and plant proteins. This has been studied from hunter-gatherers to post-agricultural societies (1), from traditional diets that enabled thriving civilizations to diets in post-industrialized countries (2, 3), and more recently, in western countries with the emerging trend toward more plant-predominant diets.

Changes in plant and animal protein intake raise classic nutritional questions. One in particular concerns the possible risk of deficiency with diets too low in animal protein, since animal protein foods contribute significantly to the intake of indispensable nutrients (4). However, plant proteins are also important for the intake of some indispensable nutrients and fiber, and are also lower in saturated fats (5). Beyond the relationship to nutrient adequacy, animal/plant proteins and their packages largely affect the metabolome and the microbiota and physiological functions that are crucial for long-term health (5-8). Accordingly, there have been many contrasting associations reported recently between plant and animal protein intake and mortality, especially regarding cardiovascular diseases (5, 9, 10).

More globally, plant (such as legumes, nuts and whole grains) and animal (such as red and processed meats) protein sources have heterogeneous relationships to nutrient adequacy (11) and to long-term health regarding cardiovascular diseases (7, 12-14) and cancers (15, 16). There is indeed a challenge for food-based dietary guidelines to point out what proportions of plant and animal protein foods should be recommended (17, 18). However, the plant to animal protein ratio remains a poor, summarizing descriptor of dietary patterns, since two diets with the same plant to animal protein ratio can actually be very different (7). There is thus a need to analyze the overall proportion of plant protein in the diet in view of the related dietary profiles and their nutritional adequacy and healthiness.

Furthermore, current interest in the proportions of plant and animal proteins in the diet also stems from their differential association with environmental pressures, in particular greenhouse gas emissions (GHGe) and land use (19-22). Altogether, the plant to animal protein ratio in the diet appears central to the sustainability of the food systems (19, 23). This has implications for dietary guidelines that aim to encompass both human and planetary health (24-26).

Here, using advanced diet modeling and optimization, we studied whether an optimal proportion of plant protein in the diet (%PP, the percentage of plant protein in total protein intake) can be identified when taking into account the reference values for nutrients and the disease burden risks for food categories. We characterized modeled diets that departed as little as possible from prevailing diets at all levels of adequate %PP values for nutrient adequacy and long-term health to identify nutritional issues (i.e., limiting nutrients) and dietary levers (i.e., effective foods). We furthermore estimated the environmental pressures associated with modeled diets along the whole range of adequate %PP values.

## Materials and Methods

### Input of dietary data

The data used for this study were extracted from the French Individual and National Study on Food Consumption Survey 3 (INCA3) conducted in 2014-2015. The INCA3 survey is a representative cross-sectional survey of the French population; its method and design have been fully described elsewhere (27). Males aged 18-64 y (*n*=564) and pre-menopausal females aged 18-54 y (*n*=561), not identified as under-reporters, were included in the present study; the final sample contained 1125 adults (Fig. S1).

Dietary data were collected by professional investigators assisted by a dietary software from three unplanned, non-consecutive, 24h dietary recalls spread over a three-week period. Portion sizes were estimated using validated photographs (27), and the nutrient contents of different food items came from the 2016 food composition database operated by the French Information Centre on Food Quality (CIQUAL) (28). Mixed foods were broken down into ingredients and then gathered into 45 food groups (Table S1). For each sex, the nutrient content of each food group was calculated as the mean nutrient content of food items constituting the food group weighted by their mean intake by the sex considered, as previously described (29). All dietary data (food group consumption and nutrient content) relate to the total population of each sex (including non-consumers).

### Multi-criteria diet optimization under constraints

Using multi-criteria optimization, we identified modeled diets (i.e., modeled consumptions of the 45 food groups) with a minimal long-term health risk and a minimal departure from the observed diet (taking into account cultural acceptability and inertia), under constraints that would ensure adequate nutrient intakes and remain within current consumption limits. In this context, we investigated the role of %PP to identify its adequate range of variations and to characterize the dietary, nutritional and environmental consequences of these variations. This non-linear optimization problem was performed using the NLP solver of the OPTMODEL procedure of SAS software version 9.4 (SAS Institute Inc., Cary, NC, USA.). Optimization was implemented at the population level but in males and females, separately. The optimized diets of males and females were then averaged to derive optimized diets for the adult population.

### Objectives

The main optimization objective was to minimize the long-term health risk of the modeled diet, as assessed by the Health Risk (*HR*) criterion. The *HR* criterion was set to target the dietary recommendations from the Global Burden of Diseases (GBD) based on epidemiological studies about the associations between consumption of different food groups and risk of chronic diseases (30). The *HR* criterion thus aimed to limit the consumption of three unhealthy food groups or categories (red meat, processed meat and sweetened beverages), while promoting that of six healthy food groups or categories (whole grain products, fruits, vegetables, legumes, nuts and seeds, and milk) until their minimum risk exposure levels (TMREL) were reached. According to the most recent (2019) estimates from the GBD, TMREL values were 0 g/d for red meat, processed meat and sweetened beverages, and 150, 325, 300, 95, 14.5 and 430 g/d, respectively, for whole grain products, fruits, vegetables, legumes, nuts and seeds, and milk (30). In our study, the *HR* criterion was thus expressed and minimized as:

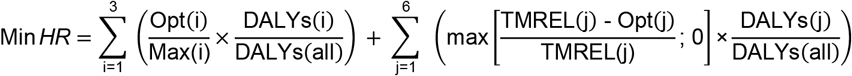

Where i denotes the food groups to be decreased (red meat, processed meat and sweetened beverages), j denotes the food groups to be increased (whole grain products, fruits, vegetables, legumes, nuts and seeds, and milk), Opt(i) and Opt(j) are the optimized consumptions of food groups i and j, respectively (in g/d), Max(i) is the upper limit of consumption of food group i (in g/d), TMREL(j) is the TMREL value of food group j (in g/d), DALYs(i) and DALYs(j) are the disability-adjusted life-years (DALYs) associated with excessive or insufficient consumptions of food groups i and j, respectively (in y), and DALYs(all) is the sum of all DALYs(i) and DALYs(j). The Max values used were the maximal recommended consumption of unhealthy foods in line with the French dietary guidelines (31): 71g/d for red meat, 25g/d for processed meat and 263 g/d (corresponding to the average portion size) for sweetened beverages intake. The TMREL and DALYs values used were issued from the most recent (2019) estimates from the GBD (30) adapted to our study context (by using sex-specific and French DALYs values, Table S2).

We also evaluated how the modeled diets deviated from current diets, in order to consider inertia to changes in food consumption, which is one way to account for social/cultural acceptability. The Diet Departure (*DD*) criterion was defined as the sum of the squares of the differences between observed and optimized food group consumption, standardized by their observed standard deviations, as previously explained (29). *DD* was thus expressed and minimized as:

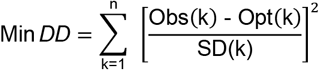

Where k is the number of food groups (n=45), Obs(k) and Opt(k) are respectively the observed and optimized consumption of food group k (in g/d) and SD(k) is the current standard deviation of the consumption of food group k.

### Constraints

During diet optimization, the total energy intake was constrained to stay within ±5% of its observed value. Thirty-five nutritional constraints were applied to ensure adequate nutrient intake in the male and female populations (Table S3), based on the most recent reference values from the French Agency for Food, Environmental and Occupational Health & Safety (ANSES) (32). We did not consider any constraints for vitamin D, because its reference value is known to be much too high to be reached by a non-fortified diet alone (29, 31). As the absorption of iron and zinc is dependent on dietary factors, the requirements were based on bioavailable iron and zinc calculated from the dietary intake using equations that predict their absorption (33-35), as detailed in a previous study by our group (29). This previous study had demonstrated that current recommendations regarding bioavailable iron and zinc are very constraining when trying to model healthier diets, these recommendations being much higher than current intakes (e.g., there is a current iron-deficiency anemia prevalence of 4.1% in French women) (29). Therefore, like in this previous study, we used threshold values lower than current reference values. They correspond to a deficiency prevalence of 5%, because such flexibility enables the identification of diets that are apparently healthier overall, with a better balance in DALYs due to less cardiometabolic disease, despite a higher prevalence of iron-deficiency anemia (29). In addition, to take into account the slightly lower digestibility of plant *vs* animal proteins regarding the nutritional constraint on protein requirement, a 5% penalty was applied to protein intake from plant protein food items, as previously described (36). As the intake of individual amino acids is generally adequate when the protein intake is sufficient in a varied diet (37), only protein requirements were considered in the model constraints, but we have *a posteriori* verified that modeled diets also contained adequate intakes of indispensable amino acids by using a database of the amino acid composition of food groups (Supporting Information Text S1).

Moreover, some acceptability constraints were applied to the food group consumption (Table S4). Acceptability constraints aimed to keep the food group intakes within the range of observed intakes, by bounding each food group intake between its 5^th^ and 95^th^ percentile of observed consumption in males and females separately. We did not do this for the unhealthy food groups or categories (red meat, processed meat and sweetened beverages), for which a dietary constraint with an upper limit was already defined according to the French dietary guidelines. Another exception was made for some healthy food groups (legumes and milk) that had 95^th^ percentile values slightly lower than TMREL values, and for which the upper limit has thus been raised to their TMREL values.

### Optimization strategy

We firstly aimed to determine the range of adequate %PP values in the diet that would ensure nutrient adequacy with a minimal long-term health risk. This first problem of identifying the adequate %PP range was addressed by optimizing the *HR* criterion under all the nutritional and acceptability constraints, with an additional constraint on %PP that was iteratively parameterized according to a grid search. This grid search constraint forced the %PP value to be equal to x%, with x% varying from 0% to 100% by steps of 5% (or even 1% at the edges of the adequate %PP range). As this problem was often non-uniquely identifiable, leading to different solutions with slightly distinct dietary patterns but similar *HR* values (especially for the intermediate %PP values that allowed for a variety of food group combinations with a similarly null *HR* value), we choose to systematically select the dietary solution that was the most acceptable *a priori*, based on the lowest departure from the current diet. According to the hierarchical method in multi-criteria optimization (38), this second problem of diet selection was addressed in a second stage. This time it was done by optimizing the *DD* criterion under the constraint that *HR* was equal to its previously identified minimal value, always under all the nutritional and acceptability constraints, and the grid search constraint on %PP covering its previously identified adequate range.

### Limiting nutrients and contribution of food groups to their intake

We conducted a dual value analysis to better characterize the tensions between %PP, nutrient adequacy and long-term health. We reported the dual values associated with the %PP equality constraint and the nutritional constraints during *HR* optimization (obtained during the first problem solving, as explained above), which represent the potential *HR* gain if the limiting bound (lower or upper) of the considered constraint was relaxed by one unit. In order to compare the relative influence of nutrients, their dual values were standardized to represent the potential *HR* gain if the limiting bound was relaxed by 10%, to classify nutrients from the most limiting (higher absolute standardized dual value) to the least limiting (lowest absolute standardized dual value). For the most limiting nutrients in the different modeled diets (i.e., nutrients with the most active constraints), we studied contributions of different food groups to intake of that particular nutrient in each modeled diet identified for each adequate %PP value (i.e., in the modeled diets resulting from the second problem solving, as explained above).

### Sensitivity analysis

We also conducted a sensitivity analysis to assess the influence of some constraints of particular interest. We thus compared the results obtained when requiring the deficiency prevalence to be ≤1% rather than ≤5% (main model) in the nutritional constraints for bioavailable iron and zinc (their alternative threshold values are given in Table S3), and when removing or not (main model) all the dietary and acceptability constraints on food group intakes.

### Diet environmental impacts

Finally, to assess environmental pressures related to the observed and modeled diets, we used the French agricultural life cycle inventory database AGRIBALYSE® v3.1; its methodological approach (summarized in Supporting Information Text S2) has been described elsewhere (39-41). In particular, we evaluated the food-related GHGe (in kg CO_2_eq, with the non-CO_2_ GHGe included and weighted according to their relative impact on warming), land use (referring to the use and transformation of land, dimensionless), water use (relating to the local scarcity of water, in m^3^ water deprivation) and fossil resource use (use of non-renewable fossil resources such as coal, oil, and gas, in MJ), together with a single environmental footprint score (dimensionless) that aggregated 16 indicators (42).

## Results

### Range of adequate %PP values and identified tensions between %PP, nutrient adequacy and long-term health

The adequate %PP range compatible with nutrient adequacy was 16%-82% in males and 16%-77% in females, and only the 25%-70% %PP range was additionally compatible with a minimal health risk (*HR* criterion) for both sexes (**Table 1**). In this narrower range, a null *HR* value was attained by the removal of unhealthy foods (red meat, processed meat and sweetened beverages) and an increase in healthy foods (whole grain products, fruits, vegetables, legumes, nuts and seeds, and milk) up to or above their TMREL values (30).

**Table 1.**
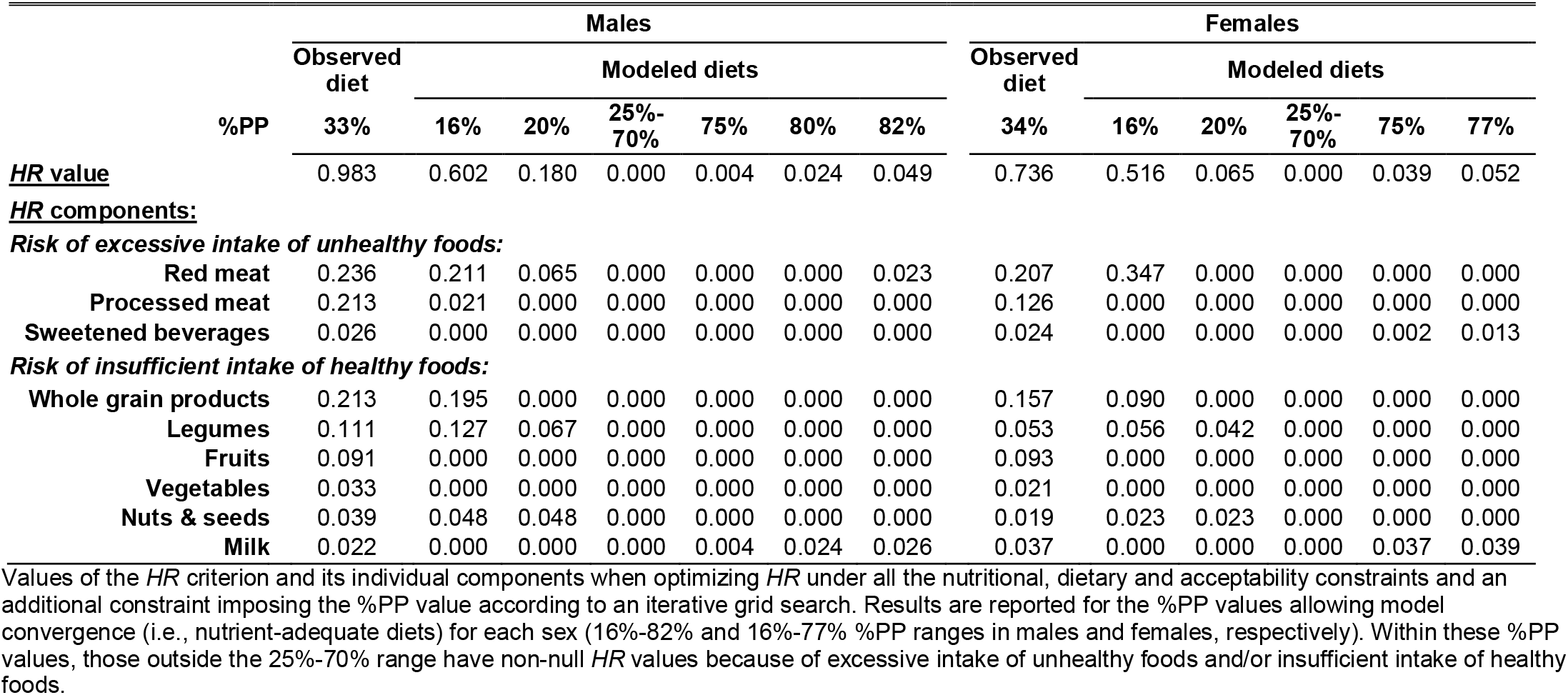
Range of adequate values of the percentage of plant protein in the diet (%PP) and corresponding minimal values of long-term health risk (*HR* criterion) in French males and females.

Among the %PP equality constraint and the nutritional constraints, none were found limiting for *HR* minimization over the 25%-70% %PP range (**Table 2**). The %PP equality constraint was limiting only for %PP values lower than 25% (strongly) and higher than 70% (more moderately, due to the lower *HR* impact of the milk decrease for the highest %PP values than of the red meat increase and whole grain product decrease for the lowest %PP values). Nutrients identified as increasingly limiting as %PP decreased below 25% were fiber, sugar (excluding lactose), saturated fatty acids and atherogenic fatty acids (lauric, myristic and palmitic acids). As %PP decreased below 25%, it was hence increasingly challenging to maintain sufficient intake of fiber and non-excessive intakes of sugar and fatty acids (as shown by the opposite sign of their dual values), which resulted in dietary solutions of increasingly degraded *HR* values. Nutrients that were identified as increasingly limiting when %PP increased above 70% were iodine, sodium, vitamin B2, calcium, EPA+DHA, vitamin A and α-linolenic acid in both sexes together with vitamin B12 in males and bioavailable iron in females. As %PP increased above 70%, it was increasingly challenging to maintain sufficient intakes of these nutrients and a non-excessive sodium intake. The other nutrients (n=20, those not shown in Table 2) were never limiting over the adequate %PP range, including, of note, protein.

**Table 2.**
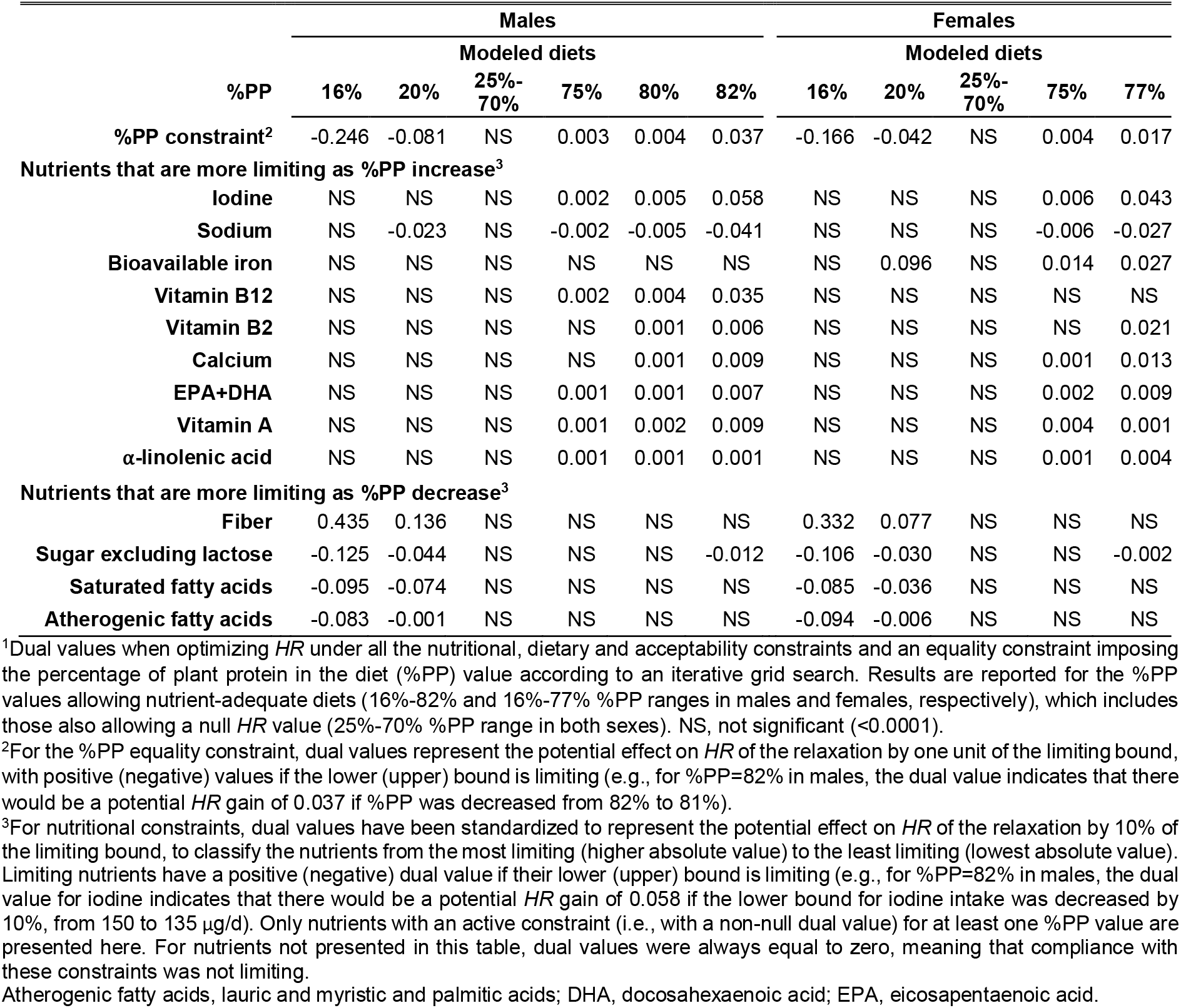
Dual values of the active constraints identified during minimization of the long-term health risk (*HR* criterion) in French males and females^1^.

Sensitivity analysis showed that being more demanding for bioavailable iron and zinc (i.e., constraining their deficiency prevalence at ≤1% rather than ≤5% as in the main model) resulted in slightly restricting the adequate %PP range on the right (16%-79% in males and 16%-70% in females), and the %PP range ensuring a null *HR* value on both sides (30%-65% in males and 35%-45% in females) (data not shown). Conversely, when suppressing all the food group consumption limits (i.e., all the dietary and acceptability constraints) from the model (Table S5), the range of adequate %PP values was expanded on both sides (8%-94% in males and 8%-92% in females), as was the %PP range ensuring a null *HR* value (16%-86% in males and 16%-84% in females), but consistently with the same limiting nutrients as in the main model (in particular, insufficient fiber intake for excessively low %PP values, or insufficient intakes of vitamin B12, iodine and EPA+DHA for excessively high %PP values).

### Modeled diets

All the modeled diets identified (**Fig. 1** and Fig. S2) were very distant from the current typical French diets, with departure values (*DD* criterion) equal to or greater than twice the standard deviation observed in the population. Furthermore, most of the modeled diets with a null *HR* value (i.e., in the 25%-70% %PP range) were all about equally distant from the observed diets, with close *DD* values (differing by less than 20%) in the 35%-65% %PP range and similar *DD* values (differing by less than 5%) in the 45%-60% %PP range.

**Fig. 1.**
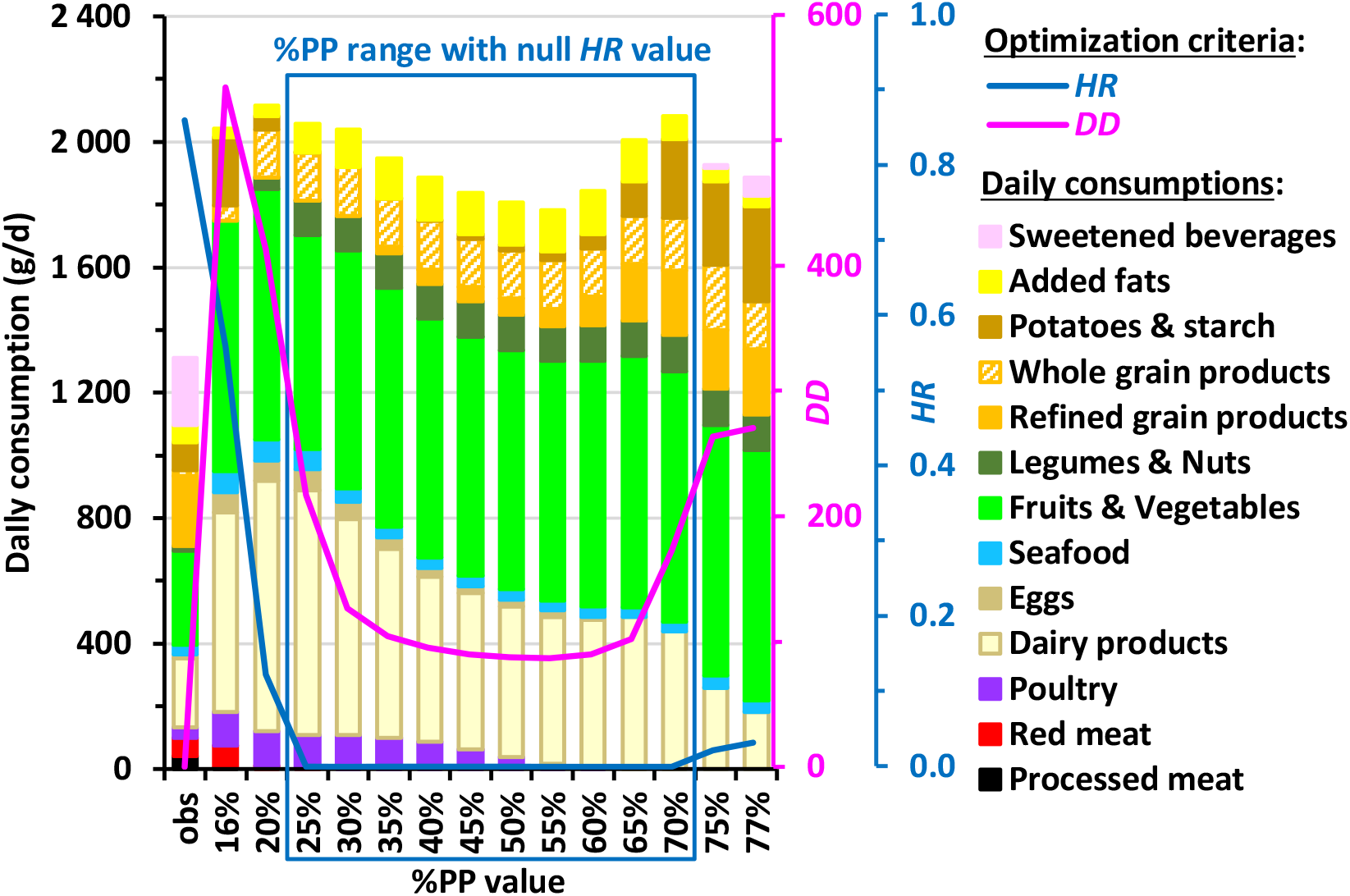
**Daily food category consumption in the observed diets (obs) and modeled diets obtained by long-term health risk (*HR*) and diet departure (*DD*) minimization under imposed percentage of plant protein in the diet (%PP) in French adults.** Results are reported for all the adequate %PP values ensuring nutrient adequacy (16%-77%), which includes those also ensuring a null *HR* value (25%-70%). The bar charts represent the cumulative consumptions of food categories (black axis on the left) and the curves represent the *HR* and *DD* values (blue and pink axes on the right, respectively). For clarity, the 45 modeled food groups are not represented here but grouped into broader categories that are included in *HR* (such as red and processed meats) or represent other protein sources (such as poultry and seafood). Consumption of water, hot beverages, alcohol and miscellaneous foods are not shown for clarity. Details about food grouping and consumptions of food categories not shown here are given in Tables S5 and S2, respectively.

Although the energy intake remained relatively stable between modeled and observed diets (by construction), the total intakes of both animal-based and plant-based foods were increased in the 25%-70% %PP range, notably owing to the important increases in milk, fruits and vegetables up to or above their TMREL values (Table S6 and Fig. S4). Regarding plant products, all the modeled diets exhibited dramatic increases in fruits and vegetables, whole grain products and legumes and nuts. Regarding animal products, red and processed meats were readily removed as %PP increased. These meats were replaced by poultry and eggs, which transiently increased, the modeled diets then being meat-free from PP%=60%. Dairy and seafood were the only remaining animal products at the right end of the adequate %PP range (Fig. S3).

Over the entire adequate %PP range, including meat-free diets, the intakes of protein and of each indispensable amino acid were always much higher than their 98% safe intake thresholds (**Fig. 2** and Table S7 and Fig. S4).

**Fig. 2.**
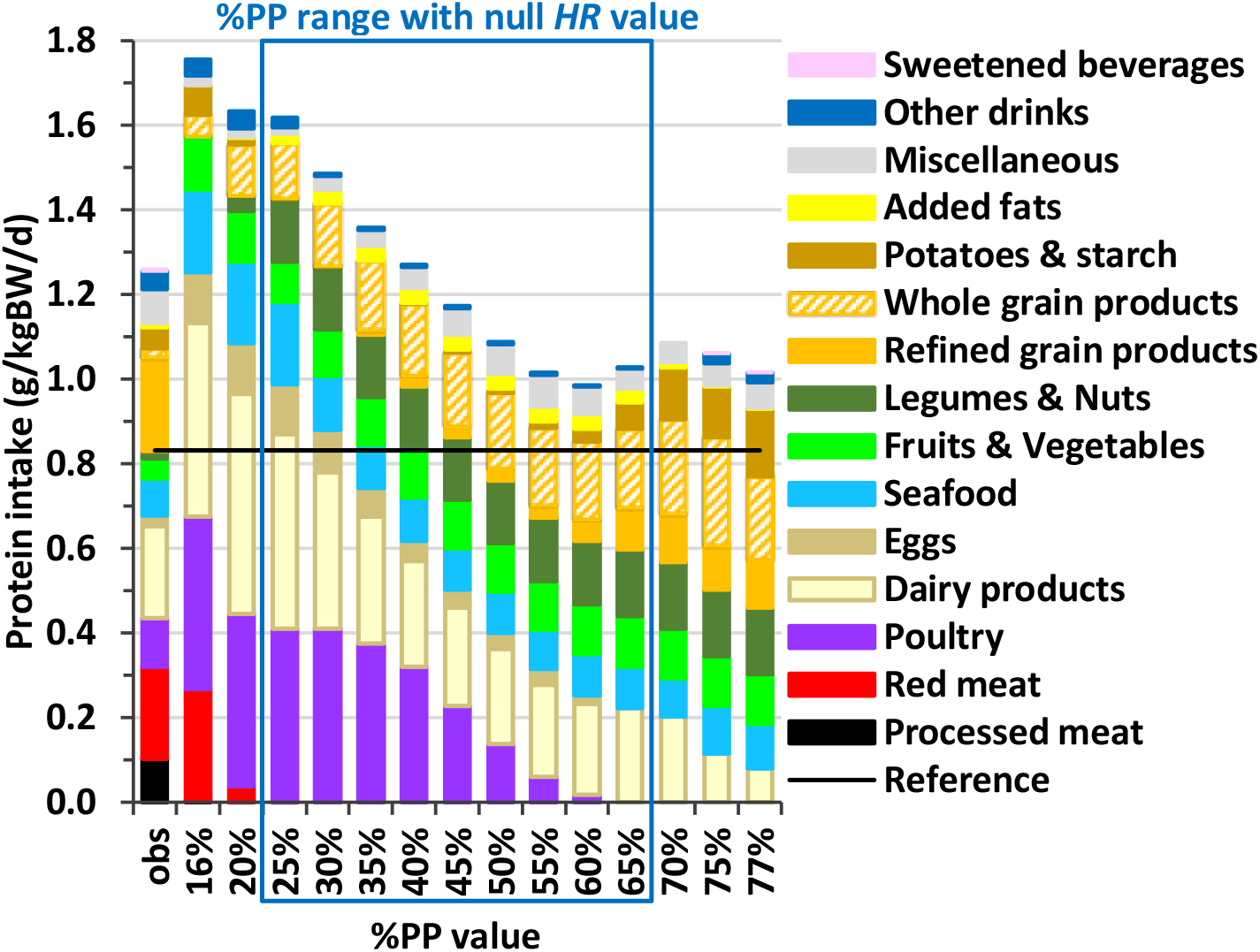
**Contribution of food categories to protein intake in the observed diets (obs) and modeled diets obtained by long-term health risk (*HR*) and diet departure minimization under imposed percentage of plant protein in the diet (%PP) in French adults.** Results are reported for all the adequate %PP values ensuring nutrient adequacy (16%-77%), which includes those ensuring also a null *HR* value (25%-70%). Sections inside the bars represent the contributions of food categories to protein intake (in g of protein/kg of BW/d). See Table S5 for the detailed composition of food categories.

### Contributions of food groups to limiting nutrient intakes

Regardless of their %PP value, all the modeled diets were nutrient-adequate, in contrast with observed diets (Table S8).

As %PP increased, it was increasingly difficult to maintain sufficient intakes of bioavailable iron, vitamins B12, B2 and A, and iodine and calcium, owing to the decreases in the animal products that were their main contributors (red meat, dairy products and eggs) (Fig. S5). The EPA+DHA and α-linolenic acid intakes, which are largely insufficient in the observed diets, were made sufficient in all the modeled diets by increases in their main contributors, respectively, seafood and added fats, with difficulties to maintain them sufficient for the highest %PP values (Fig. S5). The sodium intake, which is dramatically excessive in the observed diets, was reduced to its upper limit in all the modeled diets as a result of removing processed meat and reducing refined grain products, with difficulties to maintain sodium not excessive for the highest %PP values, due to increases in some starch and miscellaneous foods (Fig. S5). Conversely, as %PP decreased, it was increasingly difficult to maintain a sufficient intake of fiber and non-excessive intakes of sugar and saturated fatty acids, due to the meat and dairy increases (Fig. S6).

### Environmental impacts of modeled diets

Across modeled diets, GHGe gradually decreased as %PP increased until %PP=70%, where the GHGe were ∼50% lower than with the observed diet (**Fig. 3**). Similar trends were observed for land use and, to a lesser extent, fossil resource use (Fig. S7), with 40% and ∼20% decreases, respectively, from the observed to the modeled diet with %PP=70%. In contrast, water use was ∼25%-50% higher for the null-*HR* modeled diets than for the observed diet, due to their very high levels of fruits and vegetables that were by far the most water-demanding food groups (Fig. S7). Overall, at the level of the single environmental footprint score that aggregated 16 indicators, the same trend was observed as for GHGe, with a 37% decrease in this aggregated score from the observed to the modeled diet with %PP=70% (Fig. S7).

**Fig. 3.**
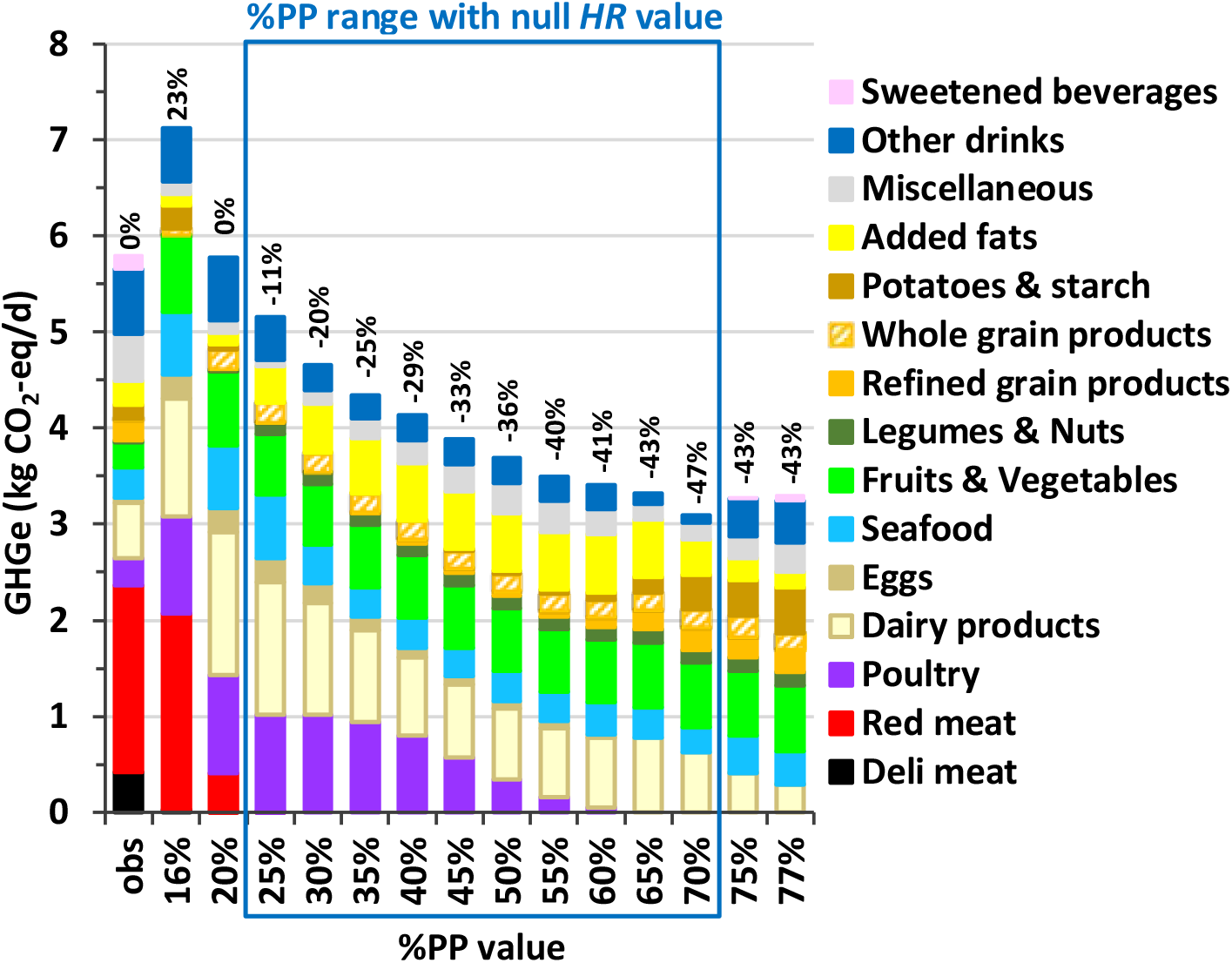
**Greenhouse gas emissions (GHGe) associated with the observed (obs) and modeled diets obtained by long-term health risk (*HR*) and diet departure minimization under imposed percentage of plant protein in the diet (%PP) in French adults.** Results are reported for all the adequate %PP values ensuring nutrient adequacy (16%-77%), which includes those ensuring also a null *HR* value (25%-70%). Sections inside the bars represent the contributions of food categories to GHGe (in kg CO_2_-eq/d), and values above the bars represent the relative deviation in GHGe from its observed value (in %). See Table S5 for the detailed composition of food categories.

## Discussion

Gathering all nutritional information over a large spectrum that covered nutrient reference values and long-term health risks, our study formally establishes ranges of plant protein proportion (%PP) for nutrient-adequate and healthy diets. One major finding is that there is no optimal %PP value, as we found a spectrum of similarly healthy diets over the 25%-70% range. However, diets in the upper end were associated with substantially lower GHGe and overall environmental impact.

A wide dietary %PP range, from ∼15% to 80%, was found compatible with providing all nutrients in adequate amounts. Our results do not agree with those of Vieux *et al*., who recently argued that %PP must be <50% to ensure nutritional adequacy (43). However, in this diet optimization study, solutions with %PP >50% were rejected not because of their true intrinsic inability to meet nutrient requirements but because of an incorrect problem formulation, as we recently pointed out (44). Furthermore, by not analyzing how the constraints considered affected the results and by not identifying the limiting nutrients, this work was not informative about the nutritional barriers to increasing %PP, which was our concern here along with its other health and environmental impacts. In our study, as shown by sensitivity analysis, the wide %PP range identified as compatible with nutritional adequacy was slightly restrained by the considered constraints for food acceptability, whereas the nutritional issues identified remained broadly the same with or without these constraints. From a nutritional viewpoint, no diet with %PP < ∼15% was able to provide enough fiber and non-excessive amounts of saturated fatty acids, while also satisfying all constraints for nutrient intakes and food acceptability. More interestingly, given the ongoing dietary transition, we could not find diets with %PP > ∼80% that would provide sufficient amounts of a large set of nutrients, particularly iodine, vitamin B12 (in males), bioavailable iron (in females), calcium and EPA+DHA. These nutrients are considered to be at issue in vegetarian diets (except calcium and iodine in lacto-ovo-vegetarian), notably calcium and B12 in predominantly plant-based diets (45, 46). From a dietary viewpoint, as shown when approaching the critical value of %PP=80%, dairy appeared to be key to preventing iodine and calcium shortages. Seafood, meanwhile, appeared critical to providing EPA+DHA (with oily fishes as the main source) as well as iodine and B12. Milk and seafood were the last remaining animal products at the highest %PP values, confirming their importance as healthy, nutrient-dense protein sources (47). Healthy plant protein sources such as legumes and nuts apparently could not replace milk or seafood. This is because they actually reached their upper allowed intake very early (as soon as %PP=25% for legumes), which indicates that they constitute an effective dietary lever. However, even when removing all food intake limits (in sensitivity analysis), it remained impossible to obtain 100% plant-based diets because of the same nutritional issues (insufficient intakes of vitamin B12, iodine and EPA+DHA). Our findings do not indicate that vegetarian (without seafood) or vegan diets (without seafood and dairy) cannot be nutritionally adequate. It means that solutions for diets that are entirely or almost entirely plant-based should rely on additional food products than those presently consumed by the general population, including fortified foods (48-50). This warrants further studies about the potential of new foods to extend the limit of the %PP range identified as adequate here.

Within the wide range of nutrient-adequate %PP values, we did not find a single optimal diet, but a large range of diets with %PP from 25% to 70%. These diets were all optimal when considering their health value, because their food consumptions complied with minimum-risk exposure levels. These consisted of no red meat and high levels of fruits and vegetables, whole grains, legumes, nuts and milk, in line with dietary guidelines (51). Modeled healthy diets were variations of this pattern, which explains why they were also similarly distant from current diets. Within this healthy pattern spectrum, the increase in %PP was predominantly related to the decrease in total and animal proteins. This occurred mostly in poultry and eggs and, to a lesser extent, dairy. This finding aligns well with the current spectrum of observed diets, with plant-based diets being higher in plant protein but especially low in total and animal protein (52, 53). This could simply be ascribed to the higher protein density in animal protein sources compared to plant protein sources. Also, the nutrients identified as limiting at the borders of the healthy %PP range appear to be related to the nutrient density of animal vs plant protein sources when expressed relative to protein density. However, the dietary protein amount was never limiting, even at the highest %PP levels. There is a growing consensus that the protein package and not the protein per se are important to the question of plant to animal protein ratio in the diet (5, 7). Likewise, indispensable amino acid amounts were well above reference values based on requirements. In real diets, composed of a mix of different types of proteins that complement each other, sufficient amounts of protein appear to guarantee sufficient amounts of amino acids (20, 53, 54).

Distance from the prevailing diets is often used in diet modeling to take into account so-called cultural acceptability (55-57), also referred to as dietary inertia (58). In this study, healthy diets in the 35-65% %PP range departed rather similarly from the prevailing diets, which are still at ∼35% %PP. This confirms that the plant to animal protein ratio is, by itself, a poor descriptor of diet characteristics, and so blanket statements about the right %PP are not warranted. Given that modeled healthy diets ranging from 35% %PP (the level of the current diets in western countries) to 65% %PP were all very distant from current diets, our study also shows that overcoming dietary inertia is required for healthy diets, irrespective of the plant to animal protein target ratio (59).

The GHGe and overall composite score for environmental pressures were lower for healthy modeled diets than observed diets, and all the more as %PP increased. A large body of literature has reported that diets which are more plant-based are associated with lower environmental pressure, and vice versa, whether diets were modeled (46, 57, 60, 61), observed (62-64) or composite (23). However, until our study, this relationship had not yet been shown according to %PP in healthy diets. In our setting, %PP was strongly associated with the environmental impact of healthy diets. As compared to the prevailing diets, lower GHGe and composite score are firstly explained by the removal of total red meat in all healthy diets, red meat accounting for ∼1/3 of the pressure in prevailing diets. This is in line with the literature that points to red meat and associated sustainability concerns (61, 65). Finally, we found that other environmental pressures (land use and fossil resource use), except water use, had similar patterns of change, in line with the literature (19, 46). The general relationship between %PP and environmental pressure can mostly be ascribed to the fact that animal sources are rich in protein, and that livestock breeding is associated with higher resource use, higher land use, and higher GHGe (21, 66-68). Nevertheless, further investigation of the relationship between %PP and environmental impacts would require prioritizing the minimization of environmental impacts over that of diet departure. Therefore, we cannot rule out the possibility that moderate %PP diets, if well-designed, may have as low environmental impacts as high %PP diets, at the cost of a larger diet departure.

This study has some limitations. We modeled diets according to changes in intakes of food groups, based on the present food repertoire and current intake levels in the population. Food grouping is critical in diet modeling (31), and food diversity and composition can change rapidly in western countries, as seen by recent changes (69). A similar limitation applies to the assessment of a diet’s environmental impacts, for which also we did not consider variations related to food production systems (70, 71). Nevertheless, we used a classical food grouping, which helps represent dietary patterns at an appropriately high level of detail. We also believe that using standard/traditional foods in modeling provides a good starting point to evaluate the situation before considering changes in the food offer or food composition. Our study uses sources of information as background parameters, including references/targets for nutrients and food categories. Clearly, there are many uncertainties in this regard (31). Nonetheless, we believe that a strength of our study is our use of a conceptual framework that aggregates most of the state of the art knowledge in nutrition.

To conclude, we identified that the range of equally optimal %PP values for nutrition and health is wide (25%-70%), and that all of these healthy diets deviate greatly from prevailing diets. From a public health perspective, there is no unique, optimal %PP value when considering nutrition and health alone. However, significant changes in current eating habits are nonetheless required to achieve healthier diets. Moreover, in the higher end of the plant protein percentage range, modeled healthy diets have a lower environmental impact and are thus more sustainable than other healthy diets. At %PP > ∼80%, changes in food repertoire diversity, food composition, nutrient enrichment or nutrient supplementation are required for fully nutrient-adequate diets.

## Data Availability

Data described in the manuscript will be made available upon request pending application and approval.

https://www.data.gouv.fr/fr/datasets/donnees-de-consommations-et-habitudes-alimentaires-de-letude-inca-3/#resources

https://www.data.gouv.fr/fr/datasets/agribalyse-r-synthese-1/#resources

## Abbreviations used

ANSES: French Agency for Food, Environmental and Occupational Health and Safety
CIQUAL: French Information Centre on Food Quality
DALYs: Disability-adjusted life-years
*DD*: Diet departure criterion
GBD: Global Burden of Diseases
GHGe: greenhouse gas emissions
*HR*: Health risk criterion
INCA3: Third Individual and National Study on Food Consumption French Survey
%PP: Percentage of plant protein in the diet
TMREL: Theoretical minimum risk exposure level

## Acknowledgments

The authors thank Vincent Colomb and Mélissa Cornélius (Ademe) for their important support in the use of the AGRIBALYSE database.

## Author Contributions

HF and FM designed the research; HF conducted the research and analyzed the data; AD, EP, FM, JW, J-FH and EK-G provided methodological support and help with interpretation of the results; HF and FM wrote the first draft of the manuscript; and all authors provided critical comments on the manuscript. HF and FM had primary responsibility for the final content, and all authors have read and approved the final manuscript.

## Data Sharing

Data described in the manuscript, code book, and analytic code will be made available upon request pending application and approval.

