## Supplemental material for "Plant to animal protein ratio in the diet: nutrient adequacy, long-term health and environmental pressure"

**This SI file includes:**

SI Text S1 to S2
Tables S1 to S8
Figures S1 to S7
SI References

**Supporting Information Text S1.** Amino acid database for food items of the third Individual and National Study on Food Consumption French Survey (INCA3).

An amino acid (AA) database was developed for the 1761 food items in the full repertoire of adults in the INCA3 study, using both the method described here and the database developed by de Gavelle *et al.* (1).

AA content databases

The AA contents of different food items were collected from published French sources (2) and international databases (3). These data came from analytical data on AA obtained using automated AA analyzers (involving ion-exchange chromatography) or high-performance liquid chromatography (HPLC).

Assignment to INCA3 food items

To assign the AA contents of the foods analyzed to INCA3 food items, we used the following procedure:

*Step 1: Direct analytical data from French published sources*
Very few analyses had been performed on French foods using chromatographic methods to analyze 18 AAs. Data from a study on the nutritional value of meat by the *Centre d'Information des Viandes* were used for most of the beef, veal, lamb, horse meat and offal food items (2).

*Step 2: Data based on "similar" food items*
When no direct analytical data corresponded to INCA3 food items, assignments were made using "similar" food items. Firstly, if data were available on a different form of the same food (*e.g.,* cooked and not raw), we hypothesized that the AA profiles of the proteins were not modified by the cooking processes, and that the data for the different forms of the same food were assigned to the INCA3 food items. Then, if data were unavailable for a food item but existed for similar species (*e.g.,* food from different cuts of the same animal), we assigned the similar food to the INCA3 food item. In order to compare AA profiles between different foods, we conducted our analyses in mg per gram of total nitrogen (mg/g of N).

*Step 3: Use of recipes to break down mixed food items*
Mixed INCA3 food items for which no data were found (*e.g.,* chili con carne or lasagna) were broken down into ingredients using the INCA3 recipe table. The food item AA content was calculated as a combination of the AA contents of its ingredients.

*Step 4: Assignment to 0 for foods containing very little or no protein*
The AA content of some INCA3 food items was assigned to 0 as these foods contained very low or no protein levels (*e.g.,* oil or alcoholic beverages).

Calculation of the AA contents of foods

We used the 2016 nutritional composition database from the French Information Centre on Food Quality (CIQUAL) (4) for protein content, which we multiplied by the AA contents (in mg/g of N) of the databases from which the data were borrowed. The CIQUAL table expresses protein content in g/100 g of food. For database matching, a conversion was therefore necessary and was carried out using the following formula: AA (mg/100 g of food) = AA (mg/g of N) * protein (g/100 g of food) / Jones' factor (N-to-protein conversion factor).

The Jones' factor used is from the USDA database (3).

**Supporting Information Text S2.** Methodological approach for the evaluation of diet environmental impacts.

The environmental pressures associated with the observed and modeled diets were estimated by indicators resulting from the matching of the 2516 food items consumed in France with those of the French database AGRIBALYSE® v3.1, a recent update of the previous 3.0.1 version developed by the French Agency for the Environment and Energy Management (ADEME) (5). In the 3.1. update, a few identified errors were corrected and some methodological improvements and enhancements have been developed. These improvements were carried out by EVEA S.A.S. Cooperative in collaboration with AGRIBALYSE® partners: GIS REVALIM, ADEME, ITERG, CIRAD, ACTALIA, ANMF, as well as the firm GINGKO21. This version has been reviewed by GIS REVALIM.

Environmental indicator estimations are based on the method of Life Cycle Assessment (LCA), the scope of which is "from field to plate". The perimeter of the indicators covers each process of the value chain (production of raw materials, transport, transformation, packaging, distribution and retailing, preparation at the consumer's place and disposal of packaging) and these processes have been split into two phases: 1) production and 2) post-farm. Of note, losses and wastes (other than the non-edible parts) at home, as well as transport from the retail to the household, have not been considered.

Overall, the method is based on the international LCA standards: ISO 14040 (6) and ISO 14044 (7), LEAP guidelines (8) and PEF (9), and the finalized indicators are provided per kg of product and are detailed per process.

For the agricultural phase of plant products, all upstream processes (notably input production) excluding storage or drying are included except for ingredients in processed food. In the case of animal products, all operations including the phases of production, transport and storage of feed, fattening of animals, milking, construction and maintenance of buildings and machinery have been considered. The scope chosen is consistent with those defined in GESTIM (10) and ecoinvent® (11).

In AGRIBALYSE® v3.0.1, the LCI (Life Cycle Inventory) data covered the period 2005-2009, except for perennial crops (2000-2010). The variety of production systems was considered by applying coefficients based on the share of systems in national production. The allocation rules are varied and are based on international recommendations as described by the ISO 14040/14044 standards (6, 7). In particular, allocations have been developed in order to distribute organic nitrogen fertilizers and mineral fertilizers (P and K) between crop sequences. Biophysical allocations were used for animal production (milk *versus* meat). The biophysical models used for animal production and allocations by type of productions are presented in the full report (12) according to the reference AFNOR-BPX 30-323 (13) and in compliance with the ISO 14044 standard (7) according to three rules in descending order: 1) avoid allocation, 2) biophysic allocation and 3) economic allocation.

A characterization method recommended by the European Commission (Environmental Footprint 3.0) translates the input and output flows of the inventory into impacts. For the background data (inputs in construction, raw materials, etc.) the ecoinvent® database is used to assess the indirect emissions (off-field emissions). The full methodology and methodological choices have been described elsewhere (12).

The transition from commodities to food as consumed introduced coefficients related to the edible part and economic allocations between co-products. The recipes were disaggregated into ingredients, and for feasibility reasons, a threshold of 95% of the ingredients covered was used. Similarly, for the origin of the ingredients, a threshold of 70% coverage was used, followed by a standardization step. The whole methodology and methodological adoptions have been described elsewhere (14) and post-farm estimations are aligned on the PEF guidelines (9).

A total of 14 midpoint indicators are available: climate change, ozone depletion, particulate matters, ionizing radiation (effect on human health), ecotoxicity, photochemical ozone formation (effect on human health), acidification, terrestrial eutrophication, freshwater eutrophication, marine eutrophication, land use, water use, Resource use, minerals and metals and Resource use, fossils. In addition, the EF 3 (environmental footprint) single score is provided, which includes these 14 indicators together with 2 further indicators related to human toxicity. The normalization and weighting factors considered in the calculation of the EF 3 score have been extensively described (9).

Concerning the indicators for food as consumed, and according to the guidelines of the PEF method (9), a quality indicator (DQR) is provided and the AGRIBALYSE® v3.0.1 database has been reviewed and criticized by RIVM and GreenDelta as well as by French agricultural and agri-food technical institutes "Peter Koch Consulting".

AGRIBALYSE® v3.1 update includes methodological changes (carbon sequestration, $N_2O$, and $NH_3$ flows, OLCA-Pest model (15), correction of land use flows, correction of nitrogen fertilizer data), the integration of new LCIs, new data on food processing (16, 17) and imported products (World Food LCA Database, v3.5), and the deployment of updates of the databases used as input, notably Ecoinvent (v3.8). All the modifications are described in a change report (18).

In particular, the synthetic environmental footprint score uses the European EF3.1 method in its unofficial interim version. The EF3.1 interim method for AGRIBALYSE® was built based on the EF 3.0 adapted method (v9.4). The major changes to the EF 3.0 adapted method concern 4 impact categories (Ecotoxicity freshwater; Human toxicity, cancer; Human toxicity, non-cancer, and Climate change).

Regarding climate change, some characterization factors of the EF3.0 method indicator have been modified based on the 6th report (AR6) of the IPCC (19). The normalization factors for the 3 impact categories Ecotoxicity freshwater; Human toxicity, cancer; Human toxicity, non-cancer have been modified based on the JRC method note EF3.1 (20).

In addition, about 50 new products, including plant substitutes, have been added. Some seasonal and out-of-season fruits and vegetables were distinguished. Out-of-season products were used for industrial products. Water use for washing fruits and vegetables was not considered except for the new products modeled (or remodeled) for AGRIBALYSE® v3.1, when they included this step. This is a limitation of the database.

**Table S1.** Food grouping: food groups and categories gathering food items consumed in the French INCA3 study.

| Food category | Food groups (Number of food items per food group) |
|---|---|
| Processed meat | Processed meats (71) |
| Red meat | Beef and veal (40)<br>Pork and other meats (39)<br>Offal (19) |
| Poultry | Poultry (24) |
| Dairy products | Milk (15)<br>Fresh natural dairy products (18)<br>Fresh sweetened dairy products (39)<br>Sweet milky desserts (22)<br>Cheeses (98) |
| Eggs | Eggs and egg-based dishes (14) |
| Seafood | Oily fishes (32)<br>Other fishes (55)<br>Mollusks and crustaceans (21) |
| Fruit & vegetables | Vegetables (149)<br>Fresh fruits (50)<br>Dried fruits (9)<br>Processed fruits: compotes and cooked fruits (13) |
| Legumes & nuts | Legumes (16)<br>Nuts, seeds and oleaginous fruits (23) |
| Refined grain products | Bread and refined bakery products (36)<br>Other refined starches (13) |
| Whole grain products | Wholemeal and semi-wholemeal bread and bakery products (15)<br>Other complete and semi-complete starches (11) |
| Potatoes & starch | Starch-based products, sweet/fat processed (61)<br>Salt/fat processed starch products (15)<br>Potatoes and other tubers (20) |
| Added fats | Animal fats and assimilated fats (4)<br>Butters and light butters (11)<br>Vegetable fats rich in alpha-linoleic acid (4)<br>Vegetable fats low in alpha-linoleic acid (24)<br>Sauces and fresh creams (55) |
| Miscellaneous foods | Sweet products or Sweet and fatty products (198)<br>Salt (6)<br>Condiments (13)<br>Aromatic herbs, Spices except salt (38)<br>Soups (30)<br>Bouillons (8)<br>Substitutes of animal products (9)<br>Other foods (14) |
| Sweetened beverages | Sweetened soda type drinks (45)<br>Fruit juices (29) |
| Other drinks | Hot drinks (22)<br>Drinking water (44)<br>Alcoholic drinks (41) |

**Table S2.** Theoretical minimum-risk exposure level (TMREL) and disability-adjusted life-years (DALYs) values used in the optimization model in French males and females.

| | | TMREL[1] (g/d) | | DALYs[2] (y) | |
|---|---|---|---|---|---|
| | | Males | Females | Males | Females |
| Unhealthy foods | Red meat | 0 | 0 | 28 562 | 20 824 |
| | Processed meat | 0 | 0 | 14 346 | 6 288 |
| | Sweetened beverages | 0 | 0 | 4 105 | 1 791 |
| Healthy foods | Whole grains | 170 | 137 | 31 405 | 10 987 |
| | Fruit | 367 | 297 | 20 130 | 9 512 |
| | Legumes | 107 | 87 | 17 103 | 3 386 |
| | Vegetables | 339 | 274 | 9 342 | 3 090 |
| | Nuts & seeds | 16 | 13 | 6 531 | 1 355 |
| | Milk | 486 | 393 | 3 521 | 2 727 |
| | Sum | | | 135 045 | 59 961 |

[1]According to the most recent (2019) estimates from the Global Burden of Diseases (GBD), the TMREL values are 0 g/d for red meat, processed meat and sweetened beverages, and 150, 325, 95, 300, 14.5 and 430 g/d, respectively, for whole cereal products, fruits, legumes, vegetables, nuts and seeds, and milk (21). As these TMREL values are overall estimates corresponding to a mean energy intake of 2300 kcal (21), we used sex-specific values adapted to the particular energy intake of males and females in our French adult population (centered around 2600 kcal and 2100 kcal in males and females, respectively).

[2]We used the most recent (2019) French sex-specific DALYs values associated with excessive/insufficient consumption of unhealthy/healthy foods, available from the Global Health Data Exchange website (http://ghdx.healthdata.org/gbd-results-tool).

**Table S3.** Nutritional constraints applied in the optimization model in French males and females[1].

| Nutrients | Units (/day) | Lower bounds[2] | | Upper bounds[3] | |
|---|---|---|---|---|---|
| | | Males | Females | Males | Females |
| Energy intake[4] | kcal | 2 470 | 1 995 | 2 730 | 2 205 |
| Retinol | µg | - | - | 3 000 | 3 000 |
| Vitamin A | µg | 750 | 650 | - | - |
| Vitamin B1 (Thiamin) | µg/kcal | 0.418 | 0.418 | - | - |
| Vitamin B2 (Riboflavin) | mg | 1.6 | 1.6 | - | - |
| Vitamin B3 (Niacin)[5] | mg NE/kcal | 0.0067 | 0.0067 | 900 | 900 |
| Vitamin B5 (Pantothenic acid)[6] | mg | 3.77 | 3.22 | - | - |
| Vitamin B6 | mg | 1.7 | 1.6 | 25 | 25 |
| Vitamin B9 (Folate) | µg | 330 | 330 | - | - |
| Vitamin B12 | µg | 4 | 4 | - | - |
| Vitamin C | mg | 110 | 110 | - | - |
| Vitamin D[7] | µg | - | - | 100 | 100 |
| Vitamin E[6] | mg | 5.28 | 4.37 | - | - |
| Vitamin K1[6] | µg | 39.47 | 34.48 | - | - |
| Calcium | mg | 950 | 950 | 2 500 | 2 500 |
| Copper [6] | mg | 1.07 | 0.89 | 5 | 5 |
| Bioavailable iron[8] | mg | 1.10 (1.40) | 1.16 (1.49) | - | - |
| Iodine | µg | 150 | 150 | 600 | 600 |
| Magnesium[6] | mg | 253.5 | 194.6 | - | - |
| Manganese[6] | mg | 1.99 | 1.52 | - | - |
| Phosphorus | mg | 550 | 550 | - | - |
| Potassium | mg | 3 500 | 3 500 | - | - |
| Selenium | µg | 70 | 70 | 300 | 300 |
| Sodium | mg | 1 500 | 1 500 | 2 300 | 2 300 |
| Bioavailable zinc[8] | mg | 2.06 (2.35) | 1.61 (1.82) | 25 | 25 |
| Water | g | 2 500 | 2 000 | - | - |
| Saturated fatty acids | % EI | - | - | 12% | 12% |
| Atherogenic fatty acids | % EI | - | - | 8% | 8% |
| Linoleic acid | % EI | 4% | 4% | - | - |
| α-linolenic acid | % EI | 1% | 1% | - | - |
| Linoleic acid / alpha-linolenic acid ratio | - | - | | 5 | 5 |
| EPA+DHA | g | 0.5 | 0.5 | - | - |
| Sugar (excluding lactose) | g | - | | 100 | 100 |
| Protein[9] | g/kg bw | 0.83 | 0.83 | 2.3 | 2.3 |
| Fiber | g | 30 | 30 | - | - |

[1]Lower and upper bounds of the nutritional constraints are based on the most recent nutrient reference values from the French Agency for Food, Environmental and Occupational Health and Safety (ANSES) (22).
[2]Lower bounds correspond to the Population Reference Intake (PRI) or lowest value of the macronutrient reference intake range. For vitamins B5, E, K1, copper, magnesium and manganese, lower bounds correspond to the 5th percentile of consumption.
[3]Upper bounds correspond to the Tolerable Upper Intake Level (UL) or highest value of the macronutrient reference intake range.
[4]Total energy intake was constrained to stay within ±5% of the current energy intake (2600 kcal for males and 2100 kcal for females).
[5]1 mg niacin equivalent (NE) is equal to 1 mg niacin or 60 mg tryptophan.
[6]For reference values that could not be based on estimated requirement (i.e. Adequate Intake), the lower bound was set at the 5th percentile of observed intake in the population.
[7]For vitamin D, a lower bound was not applied as the current reference value is too high to be reached through food intake alone.
[8]For bioavailable iron and zinc, lower bounds were not based on current reference values but on lower threshold values ensuring ≤5% deficiency prevalence. During sensitivity analysis, we also used the values shown in parentheses, which consider more demanding requirements thus ensuring ≤1% deficiency prevalence. These latter values are closer to current reference values, although still lower (but nevertheless higher than the currently observed values for bioavailable iron in females).
[9]To account for the slightly lower average digestibility of plant protein, protein intake from plants was reduced by 5% when calculating total protein intake.
Atherogenic fatty acids, lauric and myristic and palmitic acids; DHA, docosahexaenoic acid; EI, energy intake; EPA, eicosapentaenoic acid.

**Table S4.** Dietary and acceptability constraints applied for the consumption of each food group in the optimization model in French males and females.

| Food groups | Males | | | Females | | |
|---|---|---|---|---|---|---|
| | Lower consumption limit (g/d) | Prevailing diet (g/d) | Upper consumption limit (g/d) | Lower consumption limit (g/d)[1] | Prevailing diet (g/d) | Upper consumption limit (g/d) |
| Unhealthy food groups (dietary constraints)[1] | | | | | | |
| Beef and veal | 0 | 48 | 71* | 0 | 28 | 71* |
| Pork and other meats | 0 | 27 | | 0 | 13 | |
| Offal | 0 | 4 | | 0 | 1 | |
| Processed meats | 0 | 50 | 25 | 0 | 30 | 25 |
| Sweetened soda type beverages | 0 | 141 | 263* | 0 | 140 | 263* |
| Fruit juices | 0 | 80 | | 0 | 67 | |
| Other food groups (acceptability constraints)[2] | | | | | | |
| Vegetables | 20 | 176 | 400 | 18 | 160 | 387 |
| Fresh fruits | 0 | 128 | 453* | 0 | 107 | 359* |
| Dried fruits | 0 | 1 | | 0 | 1 | |
| Processed fruits: compotes and cooked fruits | 0 | 13 | | 0 | 15 | |
| Nuts, seeds and oleaginous fruits | 0 | 3 | 20 | 0 | 2 | 14 |
| Bread and refined bakery products | 28 | 168 | 354* | 19 | 115 | 316* |
| Complete and semi-complete bread and bakery products | | 11 | | | 15 | |
| Other refined starches | 0 | 98 | 276* | 0 | 72 | 188* |
| Other complete and semi-complete starches | 0 | 4 | | 0 | 4 | |
| Starch-based products, sweet/fat processed | 0 | 22 | 97 | 0 | 19 | 82 |
| Salt/fat processed starch products | 0 | 4 | 21 | 0 | 2 | 14 |
| Potatoes and other tubers | 0 | 79 | 264 | 0 | 49 | 196 |
| Legumes[3] | 0 | 13 | 107 | 0 | 6 | 87 |
| Poultry | 0 | 30 | 108 | 0 | 31 | 109 |
| Oily fishes[4] | 0 | 8 | 39* (26 for oily) | 0 | 6 | 39* (26 for oily) |
| Other fishes[4] | 0 | 22 | | 0 | 15 | |
| Mollusks and crustaceans | 0 | 5 | 28 | 0 | 4 | 26 |
| Eggs and egg-based dishes | 0 | 14 | 61 | 0 | 14 | 70 |
| Milk[3] | 0 | 84 | 486 | 0 | 75 | 393 |
| Fresh natural dairy products | 0 | 31 | 138 | 0 | 33 | 143 |
| Fresh sweetened dairy products | 0 | 50 | 179 | 0 | 48 | 168 |
| Sweet milky desserts | 0 | 19 | 93 | 0 | 16 | 73 |
| Cheeses | 0 | 49 | 131 | 0 | 36 | 94 |
| Animal fats and assimilated fats[5] | 1 | 1 | 1 | 0 | 0 | 0 |
| Butters and light butters | 0 | 10 | 33 | 0 | 10 | 30 |
| Vegetable fats rich in ALA | 0 | 0 | 32* | 0 | 0 | 32* |
| Vegetable fats low in ALA | 0 | 12 | | 0 | 10 | |
| Sauces and fresh creams | 0 | 35 | 118 | 0 | 32 | 100 |
| Sweet products or Sweet and fatty products | 9 | 103 | 251 | 9 | 83 | 215 |
| Salt | 0 | 1 | 4 | 0 | 1 | 4 |
| Condiments | 0 | 4 | 29 | 0 | 3 | 21 |
| Aromatic herbs, Spices except salt | 0 | 2 | 7 | 0 | 2 | 6 |
| Soups | 0 | 71 | 434 | 0 | 75 | 381 |
| Bouillons | 0 | 5 | 21 | 0 | 4 | 25 |
| Other foods[5] | 4 | 4 | 4 | 2 | 2 | 2 |
| Substitutes of animal products[6] | 3 | 0 | 29 | 5 | 0 | 29 |
| Drinking waters | 182 | 1 007 | - | 75 | 929 | - |
| Hot drinks[7] | 0 | 494 | 494 | 0 | 507 | 507 |
| Alcoholic drinks[7] | 0 | 216 | 216 | 0 | 59 | 59 |
| Liquids (sum of Milk, Drinking waters, Sweetened soda type drinks, Fruit juices, Hot drinks, Soups and Bouillons) | 1 061 | 2 098 | 3 777 | 738 | 1857 | 3 087 |

*These upper consumption limits correspond to coupled constraints for the groups mentioned (e.g., 71 g/d is the maximum intake for beef and veal, pork and other meats and offal grouped together).

[1]In the dietary constraints, upper bounds were applied to the food groups for which the consumption needed to be limited, in line with the French dietary guidelines (23).

[2]In the acceptability constraints, lower and upper consumption limits generally represented the 5$^{th}$ and 95$^{th}$ percentiles of consumption, respectively, of the food group in males and females, calculated using data from the third Individual and National Study on Food Consumption French Survey (INCA3), n=1125 (564 males, 561 females).

[3]For legumes and milk, the upper consumption limits in each sex were raised to their theoretical minimum-risk exposure level (TMREL) values as their 95$^{th}$ percentiles of consumption values were slightly lower (86 g/day and 43 g/day for legumes and 343 g/day and 322 g/day for milk in males and females, respectively).

[4]For fish, in order to take account of sustainable fish consumption and to limit exposure to contaminants, and in line with the French dietary guidelines on fish consumption (24), total fish consumption was limited to 39 g/day and oily fish consumption was limited to 26 g/day.

[5]For animal fats and assimilated fats and other foods, consumptions have been imposed constant and equal to the observed intakes in the modeled diets.

[6]For animal product substitutes, due to the very low value of the 95$^{th}$ percentile of consumption in men, their upper bound was set to the corresponding value in females.

[7]For hot and alcoholic drinks, the upper bound was set at the prevailing consumption.

**Table S5.** Lowest and highest adequate values of the percentage of plant protein in the diet (%PP) and corresponding limiting nutrients in modeled diets obtained by long-term health risk minimization when considering all the constraints on nutrient and food group intakes or only those on nutrient intakes in French males and females[1].

| | Lowest adequate %PP value | Highest adequate %PP value | |
|---|---|---|---|
| | | 82% in males | 77% in females |
| Main model with all the constraints on nutrient and food group intakes | 16% in both sexes<br>Fiber -<br>Sugar (excluding lactose) +<br>Saturated fatty acids +<br>Atherogenic fatty acids + | Iodine -<br>Sodium +<br>Vitamin B12 -<br>Calcium -<br>Vitamin A -<br>EPA+DHA -<br>Vitamin B2 -<br>α-linolenic acid - | Iodine -<br>Sodium +<br>Bioavailable iron -<br>Vitamin B2 -<br>Calcium -<br>EPA+DHA -<br>α-linolenic acid -<br>Vitamin A - |
| Alternative model without the constraints on food group intakes (tested during sensitivity analysis) | 8% in both sexes<br>Fiber -<br>Sugar (excluding lactose) +<br>Protein +<br>Sodium +<br>Vitamin B9 -<br>Vitamin C -<br>Manganese - | 94% in males<br>Vitamin B12 -<br>Sodium +<br>EPA+DHA -<br>Vitamin A -<br>Copper +<br>Selenium + | 92% in females<br>Iodine -<br>EPA+DHA -<br>Sodium +<br>Vitamin B12 -<br>Vitamin B2 - |

[1]In each considered case of a modeled diet with an extreme %PP value, the associated limiting nutrients (i.e., the nutrients with a non-null dual value) are listed from the most to the least limiting (i.e., from the higher to the lower absolute standardized dual value), and the limiting nutrients followed by a plus or minus sign are those being close to be excessively or insufficiently supplied, respectively (i.e., having reached their upper or lower limit, respectively).
In the alternative model (with the nutritional constraints, only, and not the dietary and acceptability constraints), limiting nutrients on a grey background are those that need to be removed from the nutritional constraints to allow model convergence for more extreme %PP values, up to diets with %PP=0% or 100%. To allow for %PP≤8%, it was thus necessary to waive nutritional adequacy in fiber for %PP≤7% and in vitamin C, sodium, manganese and protein for %PP≤1%, while to allow for %PP≥92%, it was necessary to waive nutritional adequacy in vitamin B12 and EPA+DHA for %PP≥95% and 97%, respectively, in males, and in iodine, vitamin B12 and EPA+DHA for %PP≥93%, 94% and 96%, respectively, in females.
Atherogenic fatty acids, lauric and myristic and palmitic acids; DHA, docosahexaenoic acid; EPA, eicosapentaenoic acid.

**Table S6.** Energy intake and food category consumption in observed (Obs) and modeled diets obtained by long-term health risk and diet departure minimization under imposed percentage of plant protein in the diet (%PP) in French males and females[1].

| Males | Obs diet | Modeled diets | | | | | | | | | | | | | | |
|---|---|---|---|---|---|---|---|---|---|---|---|---|---|---|---|---|
| %PP | 33% | 16% | 20% | 25% | 30% | 35% | 40% | 45% | 50% | 55% | 60% | 65% | 70% | 75% | 80% | 82% |
| Energy intake (kcal): | 2 731 | 2 470 | 2 470 | 2 486 | 2 470 | 2 470 | 2 470 | 2 470 | 2 470 | 2 470 | 2 470 | 2 470 | 2 633 | 2 730 | 2 581 | 2 603 |
| Food category consumption (g/day)[2]: | | | | | | | | | | | | | | | | |
| Processed meat | 50 | 5 | 0 | 0 | 0 | 0 | 0 | 0 | 0 | 0 | 0 | 0 | 0 | 0 | 0 | 0 |
| Red meat | 79 | 71 | 22 | 0 | 0 | 0 | 0 | 0 | 0 | 0 | 0 | 0 | 0 | 0 | 0 | 8 |
| Poultry | 30 | 108 | 108 | 108 | 108 | 108 | 103 | 72 | 49 | 30 | 10 | 0 | 0 | 0 | 0 | 0 |
| Dairy products | 233 | 678 | 854 | 840 | 808 | 695 | 590 | 569 | 539 | 505 | 508 | 544 | 486 | 404 | 42 | 0 |
| *Milk* | *84* | *486* | *486* | *486* | *486* | *486* | *486* | *486* | *486* | *486* | *486* | *486* | *486* | *404* | *42* | *0* |
| Eggs | 14 | 61 | 61 | 61 | 61 | 41 | 20 | 16 | 12 | 11 | 7 | 0 | 0 | 0 | 0 | 0 |
| Seafood | 36 | 67 | 67 | 67 | 41 | 21 | 21 | 22 | 22 | 22 | 25 | 25 | 26 | 31 | 41 | 31 |
| *Oily fishes* | *8* | *26* | *26* | *26* | *26* | *21* | *21* | *22* | *22* | *22* | *22* | *22* | *22* | *21* | *19* | *20* |
| *Other fishes* | *22* | *13* | *13* | *13* | *13* | *0* | *0* | *0* | *0* | *0* | *0* | *0* | *0* | *0* | *0* | *0* |
| *Mollusks & crustaceans* | *5* | *28* | *28* | *28* | *2* | *0* | *0* | *0* | *0* | *0* | *3* | *4* | *4* | *10* | *22* | *11* |
| Fruits & Vegetables | 318 | 854 | 854 | 721 | 804 | 827 | 838 | 843 | 845 | 843 | 839 | 854 | 854 | 854 | 854 | 798 |
| *Fruits* | *142* | *453* | *453* | *383* | *453* | *453* | *453* | *453* | *453* | *453* | *453* | *453* | *453* | *453* | *453* | *397* |
| *Vegetables* | *176* | *400* | *400* | *339* | *351* | *374* | *385* | *390* | *391* | *390* | *386* | *400* | *400* | *400* | *400* | *400* |
| Legumes & Nuts | 17 | 0 | 51 | 123 | 123 | 123 | 123 | 123 | 123 | 123 | 123 | 127 | 127 | 127 | 127 | 127 |
| *Legumes* | *13* | *0* | *51* | *107* | *107* | *107* | *107* | *107* | *107* | *107* | *107* | *107* | *107* | *107* | *107* | *107* |
| *Nuts* | *3* | *0* | *0* | *16* | *16* | *16* | *16* | *16* | *16* | *16* | *16* | *20* | *20* | *20* | *20* | *20* |
| Refined grain products | 266 | 0 | 0 | 0 | 0 | 13 | 58 | 61 | 79 | 100 | 109 | 214 | 247 | 230 | 227 | 218 |
| Whole grain products | 14 | 28 | 170 | 170 | 171 | 170 | 170 | 170 | 170 | 170 | 170 | 170 | 170 | 282 | 170 | 170 |
| Potatoes & starch | 106 | 264 | 0 | 0 | 0 | 0 | 0 | 0 | 0 | 0 | 0 | 40 | 273 | 263 | 284 | 316 |
| Added fats | 58 | 33 | 33 | 92 | 140 | 146 | 152 | 153 | 155 | 156 | 158 | 152 | 129 | 53 | 53 | 29 |
| Miscellaneous foods | 192 | 70 | 60 | 14 | 58 | 59 | 77 | 98 | 103 | 108 | 103 | 93 | 48 | 61 | 229 | 227 |
| Sweetened beverages | 221 | 0 | 0 | 0 | 0 | 0 | 0 | 0 | 0 | 0 | 0 | 0 | 0 | 0 | 0 | 0 |
| Other drinks | 1 717 | 1 372 | 1 393 | 878 | 989 | 805 | 859 | 884 | 916 | 942 | 961 | 811 | 712 | 803 | 3104 | 3 641 |
| Relative changes compared to observed diet (%)[2]: | | | | | | | | | | | | | | | | |
| Processed meat | | -90 | -100 | -100 | -100 | -100 | -100 | -100 | -100 | -100 | -100 | -100 | -100 | -100 | -100 | -100 |
| Red meat | | -10 | -72 | -100 | -100 | -100 | -100 | -100 | -100 | -100 | -100 | -100 | -100 | -100 | -100 | -90 |
| Poultry | | 256 | 256 | 256 | 256 | 256 | 237 | 136 | 62 | -1 | -68 | -100 | -100 | -100 | -100 | -100 |
| Dairy products | | 191 | 266 | 260 | 247 | 198 | 153 | 144 | 131 | 117 | 118 | 133 | 108 | 73 | -82 | -100 |
| *Milk* | | *479* | *479* | *479* | *479* | *479* | *479* | *479* | *479* | *479* | *479* | *479* | *479* | *381* | *-50* | *-100* |
| Eggs | | 336 | 336 | 336 | 336 | 190 | 42 | 16 | -15 | -20 | -51 | -100 | -100 | -100 | -100 | -100 |
| Seafood | | 87 | 87 | 87 | 14 | -41 | -40 | -40 | -39 | -38 | -31 | -29 | -27 | -13 | 15 | -13 |
| *Oily fishes* | | *209* | *209* | *209* | *209* | *148* | *152* | *156* | *159* | *160* | *160* | *156* | *159* | *155* | *124* | *143* |
| *Other fishes* | | *-42* | *-42* | *-42* | *-42* | *-100* | *-100* | *-100* | *-100* | *-100* | *-100* | *-100* | *-100* | *-100* | *-100* | *-100* |
| *Mollusks & crustaceans* | | *459* | *459* | *459* | *-69* | *-100* | *-100* | *-100* | *-100* | *-92* | *-44* | *-23* | *-11* | *96* | *350* | *117* |
| Fruits & Vegetables | | 168 | 168 | 127 | 153 | 160 | 163 | 165 | 165 | 165 | 164 | 168 | 168 | 168 | 168 | 151 |
| *Fruits* | | *219* | *219* | *169* | *219* | *219* | *218* | *218* | *219* | *219* | *218* | *219* | *219* | *219* | *219* | *179* |
| *Vegetables* | | *128* | *128* | *93* | *99* | *113* | *119* | *122* | *123* | *122* | *120* | *128* | *128* | *128* | *128* | *128* |
| Legumes & Nuts | | -100 | 206 | 643 | 646 | 644 | 644 | 644 | 644 | 644 | 644 | 667 | 668 | 668 | 668 | 668 |
| *Legumes* | | *-100* | *276* | *696* | *696* | *696* | *696* | *696* | *696* | *696* | *696* | *696* | *696* | *696* | *696* | *696* |
| *Nuts* | | *-100* | *-100* | *414* | *433* | *419* | *418* | *418* | *418* | *422* | *419* | *545* | *548* | *548* | *548* | *548* |
| Refined grain products | | -100 | -100 | -100 | -100 | -95 | -78 | -77 | -70 | -63 | -59 | -19 | -7 | -13 | -14 | -18 |
| Whole grain products | | 91 | 1 075 | 1 076 | 1 081 | 1 075 | 1 075 | 1 075 | 1 075 | 1075 | 1 075 | 1 075 | 1 075 | 1 846 | 1 075 | 1 077 |
| Potatoes & starch | | 150 | -100 | -100 | -100 | -100 | -100 | -100 | -100 | -100 | -100 | -62 | 158 | 149 | 169 | 199 |
| Added fats | | -43 | -43 | 60 | 142 | 153 | 163 | 164 | 168 | 171 | 174 | 163 | 124 | -8 | -9 | -49 |
| Miscellaneous foods | | -64 | -69 | -93 | -70 | -70 | -60 | -49 | -46 | -44 | -47 | -52 | -75 | -68 | 19 | 18 |
| Sweetened beverages | | -100 | -100 | -100 | -100 | -100 | -100 | -100 | -100 | -100 | -100 | -100 | -100 | -100 | -100 | -100 |
| Other drinks | | -20 | -19 | -49 | -42 | -53 | -50 | -49 | -47 | -45 | -44 | -53 | -59 | -53 | 81 | 112 |

| Females | Obs diet | Modeled diets | | | | | | | | | | | | | |
|---|---|---|---|---|---|---|---|---|---|---|---|---|---|---|---|
| %PP | 34% | 16% | 20% | 25% | 30% | 35% | 40% | 45% | 50% | 55% | 60% | 65% | 70% | 75% | 77% |
| Energy intake (kcal): | 2 024 | 2 205 | 2 110 | 2 042 | 1 995 | 1 995 | 1 995 | 1 995 | 1 995 | 1 995 | 2 092 | 2 205 | 2 205 | 2 205 | 2 205 |
| Food category consumption (g/day)[2]: | | | | | | | | | | | | | | | |
| Processed meat | 30 | 0 | 0 | 0 | 0 | 0 | 0 | 0 | 0 | 0 | 0 | 0 | 0 | 0 | 0 |
| Red meat | 42 | 71 | 0 | 0 | 0 | 0 | 0 | 0 | 0 | 0 | 0 | 0 | 0 | 0 | 0 |
| Poultry | 31 | 109 | 109 | 109 | 109 | 93 | 70 | 51 | 26 | 4 | 0 | 0 | 0 | 0 | 0 |
| Dairy products | 208 | 592 | 745 | 720 | 571 | 507 | 465 | 429 | 425 | 431 | 432 | 423 | 393 | 116 | 86 |
| *Milk* | *75* | *393* | *393* | *393* | *393* | *393* | *393* | *393* | *393* | *393* | *393* | *393* | *393* | *77* | *56* |
| Eggs | 14 | 70 | 70 | 70 | 48 | 34 | 32 | 29 | 28 | 28 | 13 | 0 | 0 | 0 | 0 |
| Seafood | 25 | 65 | 65 | 65 | 45 | 45 | 46 | 43 | 44 | 42 | 43 | 41 | 34 | 46 | 41 |
| *Oily fishes* | *6* | *26* | *26* | *25* | *20* | *21* | *21* | *21* | *21* | *22* | *22* | *22* | *24* | *20* | *21* |
| *Other fishes* | *15* | *13* | *13* | *14* | *19* | *18* | *16* | *10* | *6* | *0* | *0* | *0* | *0* | *0* | *0* |
| *Mollusks & crustaceans* | *4* | *26* | *26* | *26* | *6* | *6* | *9* | *12* | *17* | *20* | *21* | *19* | *10* | *11* | *20* |
| Fruits & Vegetables | 283 | 746 | 746 | 641 | 710 | 696 | 688 | 684 | 684 | 684 | 730 | 746 | 746 | 746 | 746 |
| *Fruits* | *124* | *359* | *359* | *359* | *359* | *315* | *301* | *297* | *297* | *297* | *343* | *359* | *359* | *359* | *359* |
| *Vegetables* | *160* | *387* | *387* | *282* | *351* | *381* | *387* | *387* | *387* | *387* | *387* | *387* | *387* | *387* | *387* |
| Legumes & Nuts | 8 | 0 | 22 | 100 | 100 | 100 | 100 | 100 | 100 | 100 | 101 | 101 | 101 | 101 | 101 |
| *Legumes* | *6* | *0* | *22* | *87* | *87* | *87* | *87* | *87* | *87* | *87* | *87* | *87* | *87* | *87* | *87* |
| *Nuts* | *2* | *0* | *0* | *13* | *13* | *13* | *13* | *13* | *13* | *13* | *14* | *14* | *14* | *14* | *14* |
| Refined grain products | 186 | 0 | 0 | 0 | 0 | 25 | 33 | 33 | 25 | 13 | 74 | 147 | 165 | 145 | 138 |
| Whole grain products | 19 | 70 | 137 | 137 | 137 | 137 | 137 | 137 | 137 | 137 | 137 | 137 | 170 | 137 | 137 |
| Potatoes & starch | 71 | 169 | 85 | 0 | 0 | 0 | 11 | 28 | 39 | 56 | 94 | 178 | 224 | 267 | 278 |
| Added fats | 53 | 25 | 31 | 89 | 100 | 110 | 113 | 113 | 111 | 108 | 112 | 110 | 22 | 32 | 18 |
| Miscellaneous foods | 174 | 39 | 39 | 29 | 44 | 61 | 78 | 92 | 107 | 115 | 88 | 46 | 151 | 224 | 247 |
| Sweetened beverages | 208 | 0 | 0 | 0 | 0 | 0 | 0 | 0 | 0 | 0 | 0 | 0 | 0 | 20 | 116 |
| Other drinks | 1 495 | 1 433 | 1 941 | 1 386 | 810 | 836 | 887 | 870 | 914 | 861 | 793 | 595 | 500 | 2 523 | 2 749 |
| Relative changes compared to observed diet (%)[2]: | | | | | | | | | | | | | | | |
| Processed meat | | -100 | -100 | -100 | -100 | -100 | -100 | -100 | -100 | -100 | -100 | -100 | -100 | -100 | -100 |
| Red meat | | 67 | -100 | -100 | -100 | -100 | -100 | -100 | -100 | -100 | -100 | -100 | -100 | -100 | -100 |
| Poultry | | 248 | 248 | 248 | 248 | 196 | 123 | 62 | -18 | -86 | -100 | -100 | -100 | -100 | -100 |
| Dairy products | | 185 | 258 | 246 | 174 | 144 | 123 | 106 | 104 | 107 | 108 | 103 | 89 | -44 | -59 |
| *Milk* | | *421* | *421* | *421* | *421* | *421* | *421* | *421* | *421* | *421* | *421* | *421* | *421* | *2* | *-26* |
| Eggs | | 410 | 410 | 409 | 251 | 149 | 133 | 114 | 101 | 102 | -5 | -100 | -100 | -100 | -100 |
| Seafood | | 160 | 160 | 160 | 81 | 80 | 83 | 70 | 75 | 67 | 71 | 65 | 35 | 84 | 64 |
| *Oily fishes* | | *331* | *331* | *319* | *233* | *241* | *240* | *252* | *250* | *258* | *258* | *266* | *295* | *316* | *251* |
| *Other fishes* | | *-16* | *-16* | *-11* | *23* | *19* | *7* | *-38* | *-59* | *-100* | *-100* | *-100* | *-100* | *-100* | *-100* |
| *Mollusks & crustaceans* | | *606* | *606* | *606* | *73* | *71* | *145* | *222* | *346* | *445* | *472* | *419* | *171* | *196* | *440* |
| Fruits & Vegetables | | 163 | 163 | 126 | 151 | 146 | 143 | 141 | 141 | 141 | 158 | 163 | 163 | 163 | 163 |
| *Fruits* | | *191* | *191* | *191* | *191* | *155* | *144* | *140* | *141* | *140* | *178* | *191* | *191* | *191* | *191* |
| *Vegetables* | | *142* | *142* | *76* | *120* | *138* | *142* | *142* | *142* | *142* | *142* | *142* | *142* | *142* | *142* |
| Legumes & Nuts | | -100 | 173 | 1 131 | 1 131 | 1 131 | 1 131 | 1 133 | 1 131 | 1 131 | 1 147 | 1 147 | 1 147 | 1 147 | 1 147 |
| *Legumes* | | *-100* | *276* | *1374* | *1374* | *1374* | *1374* | *1374* | *1374* | *1374* | *1374* | *1374* | *1374* | *1374* | *1374* |
| *Nuts* | | *-100* | *-100* | *486* | *486* | *485* | *485* | *493* | *487* | *486* | *543* | *543* | *543* | *543* | *543* |
| Refined grain products | | -100 | -100 | -100 | -100 | -87 | -82 | -82 | -87 | -93 | -61 | -21 | -12 | -22 | -26 |
| Whole grain products | | 262 | 611 | 611 | 611 | 611 | 611 | 611 | 611 | 611 | 611 | 611 | 779 | 611 | 611 |
| Potatoes & starch | | 139 | 21 | -100 | -100 | -100 | -85 | -61 | -44 | -21 | 32 | 151 | 216 | 278 | 294 |
| Added fats | | -52 | -41 | 69 | 91 | 109 | 116 | 115 | 111 | 106 | 113 | 110 | -59 | -40 | -65 |
| Miscellaneous foods | | -78 | -77 | -83 | -75 | -65 | -55 | -47 | -39 | -34 | -49 | -73 | -13 | 29 | 42 |
| Sweetened beverages | | -100 | -100 | -100 | -100 | -100 | -100 | -100 | -100 | -100 | -100 | -100 | -100 | -91 | -44 |
| Other drinks | | -4 | 30 | -7 | -46 | -44 | -41 | -42 | -39 | -42 | -47 | -60 | -67 | 69 | 84 |

[1]Observed data from the third Individual and National Study on Food Consumption French Survey (INCA3), n=1125 (564 males, 561 females).
[2] For clarity, the 45 modeled food groups are not represented here but grouped into broader categories (see Supplemental Table 1 for details about food grouping). Consumption data on a black background are those having been zeroed, and consumption data on a grey background are those having been raised to their maximal values, as set in the acceptability constraints (see Supplemental Table 4 for details about these consumption limits).

**Table S7**. Protein and amino acid intakes in observed (Obs) and modeled diets obtained by long-term health risk and diet departure minimization under the imposed percentage of plant protein in the diet (%PP) in French adults[1].

| | Obs diet | Modeled diets | | | | | | | | | | | | | | EAR[3] | 98% safe intake (=EAR +2*SD) |
|---|---|---|---|---|---|---|---|---|---|---|---|---|---|---|---|---|---|
| %PP | 33% | 16% | 20% | 25% | 30% | 35% | 40% | 45% | 50% | 55% | 60% | 65% | 70% | 75% | 77% | | |
| Intakes (mg·kgBW$^{-1}$·d$^{-1}$)[2]: | | | | | | | | | | | | | | | | | |
| Aspartic acid + asparagine | 109 | 176 | 157 | 155 | 141 | 128 | 120 | 111 | 102 | 95 | 93 | 96 | 105 | 106 | 104 | - | - |
| Alanine | 60 | 88 | 79 | 78 | 70 | 64 | 59 | 53 | 48 | 43 | 41 | 42 | 43 | 44 | 43 | - | - |
| Arginine | 68 | 98 | 89 | 92 | 84 | 76 | 71 | 65 | 59 | 54 | 52 | 54 | 56 | 59 | 58 | - | - |
| Methionine + Cysteine | 49 | 67 | 60 | 58 | 52 | 47 | 44 | 40 | 36 | 33 | 32 | 33 | 35 | 35 | 33 | 15 | 19 |
| *Methionine* | *31* | *46* | *41* | *39* | *34* | *30* | *27* | *24* | *22* | *20* | *18* | *18* | *19* | *18* | *17* | *-* | *-* |
| *Cysteine* | *18* | *20* | *19* | *19* | *18* | *17* | *16* | *15* | *14* | *14* | *14* | *14* | *16* | *17* | *16* | *-* | *-* |
| Glutamic acid + glutamine | 257 | 306 | 303 | 304 | 284 | 268 | 256 | 242 | 230 | 220 | 219 | 233 | 249 | 244 | 231 | - | - |
| Glycine | 50 | 71 | 62 | 63 | 57 | 52 | 50 | 45 | 41 | 38 | 36 | 38 | 39 | 41 | 40 | - | - |
| Histidine | 36 | 50 | 47 | 46 | 42 | 38 | 35 | 31 | 28 | 26 | 25 | 25 | 26 | 25 | 24 | 10 | 12 |
| Isoleucine | 55 | 80 | 76 | 75 | 67 | 60 | 55 | 50 | 46 | 43 | 41 | 42 | 43 | 42 | 40 | 20 | 25 |
| Leucine | 98 | 141 | 134 | 132 | 119 | 107 | 99 | 90 | 82 | 76 | 73 | 75 | 78 | 75 | 72 | 39 | 48 |
| Lysine | 83 | 135 | 125 | 121 | 108 | 96 | 86 | 76 | 67 | 59 | 55 | 54 | 55 | 53 | 50 | 30 | 37 |
| Phenylalanine + Tyrosine | 99 | 145 | 138 | 135 | 120 | 108 | 100 | 92 | 85 | 79 | 76 | 79 | 82 | 79 | 75 | 25 | 31 |
| *Phenylalanine* | *56* | *78* | *74* | *74* | *67* | *61* | *57* | *53* | *49* | *47* | *45* | *47* | *50* | *49* | *47* | *-* | *-* |
| *Tyrosine* | *43* | *67* | *64* | *61* | *53* | *47* | *43* | *39* | *35* | *32* | *31* | *31* | *32* | *30* | *29* | *-* | *-* |
| Proline | 88 | 108 | 109 | 106 | 96 | 89 | 84 | 80 | 76 | 74 | 74 | 78 | 82 | 78 | 73 | - | - |
| Serine | 58 | 84 | 81 | 81 | 73 | 66 | 61 | 57 | 53 | 51 | 49 | 50 | 52 | 51 | 48 | - | - |
| Threonine | 50 | 74 | 68 | 66 | 59 | 53 | 49 | 45 | 41 | 37 | 36 | 37 | 38 | 37 | 36 | 15 | 19 |
| Tryptophan | 15 | 21 | 21 | 20 | 18 | 16 | 15 | 14 | 13 | 12 | 11 | 12 | 12 | 12 | 12 | 4 | 5 |
| Valine | 65 | 95 | 90 | 88 | 78 | 70 | 65 | 60 | 55 | 51 | 49 | 51 | 53 | 52 | 49 | 26 | 32 |
| Protein | 1 240 | 1 742 | 1 617 | 1 598 | 1 462 | 1 334 | 1 245 | 1 145 | 1 060 | 987 | 956 | 993 | 1 048 | 1 020 | 980 | 660 | 818 |
| Deviations from the 98% safe intake threshold (%)[4]: | | | | | | | | | | | | | | | | | |
| Methionine + Cysteine | 157% | 252% | 214% | 203% | 174% | 149% | 130% | 109% | 91% | 76% | **68%** | 72% | 83% | 83% | 74% | | |
| Histidine | 199% | 313% | 288% | 281% | 246% | 213% | 189% | 162% | 137% | 116% | 107% | 112% | 119% | 112% | **101%** | | |
| Isoleucine | 122% | 221% | 205% | 199% | 168% | 141% | 122% | 102% | 85% | 70% | 63% | 68% | 74% | 69% | **61%** | | |
| Leucine | 105% | 194% | 179% | 175% | 148% | 123% | 106% | 87% | 72% | 58% | 52% | 57% | 63% | 57% | **50%** | | |
| Lysine | 124% | 265% | 238% | 227% | 192% | 158% | 133% | 106% | 82% | 61% | 49% | 47% | 50% | 43% | **35%** | | |
| Phenylalanine + Tyrosine | 220% | 368% | 345% | 336% | 288% | 249% | 222% | 196% | 174% | 155% | 146% | 154% | 163% | 154% | **143%** | | |
| Threonine | 163% | 289% | 258% | 247% | 212% | 181% | 160% | 135% | 114% | 96% | **87%** | 92% | 100% | 97% | 90% | | |
| Tryptophan | 206% | 330% | 313% | 303% | 258% | 224% | 201% | 176% | 155% | 137% | **130%** | 138% | 148% | 148% | 136% | | |
| Valine | 102% | 195% | 180% | 174% | 145% | 120% | 103% | 86% | 72% | 60% | 55% | 60% | 67% | 62% | **54**% | | |
| Protein | 52% | 113% | 98% | 95% | 79% | 63% | 52% | 40% | 30% | 21% | **17**% | 21% | 28% | 25% | 20% | | |

[1]Observed data from the third Individual and National Study on Food Consumption French Survey (INCA3), n=1125 (564 males, 561 females). Observed and modeled diets in French adults (mean of males and females).

[2]To account for the slightly lower average digestibility of plant protein, protein intake from plants was reduced by 5% when calculating total protein and amino acid intakes (see the Methods section).

[3]EAR, Estimated average requirement of adults for protein and indispensable amino acids, based on the Joint FAO/WHO/UNU Expert Consultation on Protein and Amino Acid Requirements in Human Nutrition (2002) (25).

[4]Over all the observed and modeled diets, individual indispensable amino acid and protein intakes were always higher than the corresponding 98% safe intake, the minimum deviation from this threshold value being indicated in bold.

**Table S8**. Daily intakes of energy and nutrients in observed (Obs) and modeled diets obtained by long-term health risk and diet departure minimization under imposed percentage of plant protein in the diet (%PP) in French males and females[1,2].

**Males**

| | Units (/day) | Obs diet | Modeled diets | | | | | | | | | | | | | | |
|---|---|---|---|---|---|---|---|---|---|---|---|---|---|---|---|---|---|
| | %PP | 33% | 16% | 20% | 25% | 30% | 35% | 40% | 45% | 50% | 55% | 60% | 65% | 70% | 75% | 80% | 82% |
| Energy intake | kcal | 2 731 | 2 470 | 2 470 | 2 486 | 2 470 | 2 470 | 2 470 | 2 470 | 2 470 | 2 470 | 2 470 | 2 470 | 2 633 | 2 730 | 2 581 | 2 603 |
| Vitamin A | µg | 805 | 815 | 841 | 764 | 750 | 750 | 750 | 750 | 750 | 750 | 750 | 750 | 750 | 750 | 750 | 750 |
| Vitamin B1 | µg/kcal | 0.54 | 0.59 | 0.63 | 0.61 | 0.67 | 0.64 | 0.62 | 0.60 | 0.59 | 0.57 | 0.56 | 0.58 | 0.61 | 0.59 | 0.56 | 0.57 |
| Vitamin B2 | mg | 2.17 | 3.20 | 3.55 | 3.14 | 2.92 | 2.58 | 2.28 | 2.17 | 2.07 | 1.95 | 1.91 | 1.84 | 1.80 | 1.70 | 1.60 | 1.60 |
| Vitamin B3[3] | mg NE/kcal | 0.016 | 0.021 | 0.019 | 0.017 | 0.017 | 0.016 | 0.015 | 0.014 | 0.013 | 0.011 | 0.010 | 0.011 | 0.012 | 0.012 | 0.013 | 0.013 |
| Vitamin B5 | mg | 6.9 | 9.6 | 9.9 | 9.2 | 9.4 | 8.7 | 8.1 | 7.6 | 7.2 | 6.9 | 6.6 | 6.6 | 7.5 | 7.4 | 6.6 | 6.8 |
| Vitamin B6 | mg | 2.1 | 3.0 | 2.3 | 2.2 | 2.4 | 2.3 | 2.2 | 2.1 | 2.0 | 1.9 | 1.8 | 1.9 | 2.5 | 2.5 | 2.4 | 2.4 |
| Vitamin B9 | µg | 347 | 438 | 496 | 503 | 548 | 524 | 511 | 510 | 506 | 503 | 498 | 527 | 563 | 565 | 543 | 534 |
| Vitamin B12 | µg | 6.2 | 10.6 | 9.8 | 9.2 | 6.4 | 5.1 | 4.7 | 4.4 | 4.2 | 4.0 | 4.0 | 4.0 | 4.0 | 4.0 | 4.0 | 4.0 |
| Vitamin C | mg | 98 | 158 | 146 | 115 | 154 | 156 | 157 | 158 | 158 | 158 | 158 | 163 | 182 | 178 | 177 | 150 |
| Vitamin D | µg | 3.5 | 5.4 | 6.4 | 6.6 | 6.7 | 5.4 | 4.7 | 4.5 | 4.3 | 4.1 | 3.9 | 3.8 | 3.5 | 2.9 | 2.1 | 1.8 |
| Vitamin E | mg | 13 | 18 | 18 | 18 | 19 | 19 | 19 | 19 | 19 | 20 | 20 | 19 | 19 | 16 | 18 | 17 |
| Vitamin K1 | µg | 118 | 278 | 263 | 238 | 205 | 226 | 230 | 232 | 234 | 236 | 241 | 251 | 256 | 249 | 267 | 305 |
| Calcium | mg | 1 065 | 1 618 | 1 817 | 1 646 | 1 495 | 1 316 | 1 210 | 1 199 | 1 172 | 1 143 | 1 147 | 1 204 | 1 150 | 1 084 | 950 | 950 |
| Copper | mg | 2.1 | 2.0 | 2.0 | 2.0 | 2.0 | 2.0 | 2.0 | 2.0 | 2.0 | 2.0 | 2.0 | 2.2 | 2.3 | 2.6 | 2.8 | 2.9 |
| Bioavailable iron[4] | mg | 1.68 | 1.80 | 1.37 | 1.25 | 1.23 | 1.24 | 1.19 | 1.16 | 1.10 | 1.10 | 1.10 | 1.10 | 1.10 | 1.19 | 1.14 | 1.19 |
| *Oral iron* | *mg* | *13* | *14* | *13* | *12* | *12* | *12* | *12* | *12* | *12* | *12* | *12* | *13* | *13* | *14* | *15* | *16* |
| *Iron absorption* | *%* | *13%* | *13%* | *10%* | *10%* | *10%* | *10%* | *10%* | *10%* | *9%* | *9%* | *9%* | *9%* | *9%* | *9%* | *8%* | *8%* |
| Iodine | µg | 175 | 253 | 265 | 249 | 226 | 190 | 171 | 167 | 163 | 159 | 156 | 164 | 165 | 150 | 150 | 150 |
| Magnesium | mg | 409 | 487 | 527 | 457 | 480 | 463 | 462 | 455 | 453 | 446 | 445 | 471 | 495 | 531 | 572 | 560 |
| Manganese | mg | 3.6 | 2.9 | 4.0 | 3.5 | 4.0 | 4.3 | 4.5 | 4.6 | 4.7 | 4.8 | 4.9 | 5.3 | 5.6 | 6.7 | 6.0 | 5.9 |
| Phosphorus | mg | 1 483 | 1 961 | 2 061 | 2 041 | 1 942 | 1 785 | 1 689 | 1 615 | 1 547 | 1 490 | 1 444 | 1 514 | 1 600 | 1 664 | 1 241 | 1 214 |
| Potassium | mg | 3 736 | 5 917 | 4 922 | 4 334 | 4 686 | 4 558 | 4 454 | 4 357 | 4 290 | 4 187 | 4 152 | 4 350 | 5 226 | 5 154 | 5 026 | 4 780 |
| Selenium | µg | 146 | 178 | 179 | 164 | 152 | 133 | 135 | 132 | 130 | 129 | 128 | 138 | 141 | 139 | 192 | 211 |
| Sodium | mg | 3 938 | 2 300 | 2 300 | 2 294 | 2 300 | 2 300 | 2 300 | 2 300 | 2 300 | 2 300 | 2 300 | 2 300 | 2 300 | 2 300 | 2 300 | 2 300 |
| Bioavailable zinc[4] | mg | 3.77 | 4.47 | 4.39 | 4.07 | 3.71 | 3.37 | 3.15 | 2.99 | 2.84 | 2.71 | 2.62 | 2.67 | 2.61 | 2.62 | 2.32 | 2.18 |
| *Oral zinc* | *mg* | *12.1* | *15.1* | *14.2* | *12.9* | *11.1* | *10.3* | *9.8* | *9.3* | *9.0* | *8.7* | *8.5* | *9.0* | *9.2* | *10.0* | *9.1* | *8.8* |
| *Zinc absorption* | *%* | *31%* | *30%* | *31%* | *32%* | *33%* | *33%* | *32%* | *32%* | *32%* | *31%* | *31%* | *30%* | *28%* | *26%* | *25%* | *25%* |
| Water | g | 2 780 | 3 099 | 3 163 | 2 564 | 2 797 | 2 500 | 2 500 | 2 500 | 2 501 | 2 500 | 2 500 | 2 500 | 2 500 | 2 500 | 4 569 | 4 988 |
| Saturated fatty acids | % EI | 14% | 12% | 12% | 12% | 12% | 12% | 12% | 12% | 12% | 12% | 12% | 12% | 11% | 10% | 9% | 6% |
| Atherogenic fatty acids | % EI | 8% | 8% | 8% | 8% | 7% | 7% | 7% | 7% | 7% | 7% | 7% | 7% | 7% | 6% | 6% | 3% |
| Linoleic acid | % EI | 3% | 5% | 4% | 5% | 5% | 5% | 5% | 5% | 5% | 5% | 5% | 5% | 5% | 4% | 5% | 5% |
| α-linolenic acid | % EI | 0.0% | 1.0% | 1.0% | 1.0% | 1.0% | 1.0% | 1.0% | 1.0% | 1.0% | 1.0% | 1.0% | 1.0% | 1.0% | 1.0% | 1.0% | 1.0% |
| Linoleic / α-linolenic | - | 8 | 4 | 4 | 5 | 5 | 5 | 5 | 5 | 5 | 5 | 5 | 5 | 5 | 4 | 5 | 5 |
| EPA+DHA | g | 0.33 | 0.71 | 0.72 | 0.73 | 0.66 | 0.50 | 0.50 | 0.50 | 0.50 | 0.50 | 0.50 | 0.50 | 0.50 | 0.50 | 0.50 | 0.50 |
| Sugar (excluding lactose) | g | 102 | 100 | 100 | 98 | 98 | 100 | 96 | 100 | 100 | 100 | 100 | 98 | 75 | 73 | 84 | 100 |
| Protein[5] | g | 103 | 126 | 123 | 119 | 110 | 100 | 95 | 87 | 81 | 76 | 70 | 73 | 78 | 83 | 69 | 68 |
| Fiber | g | 23 | 30 | 30 | 30 | 33 | 35 | 37 | 37 | 38 | 39 | 40 | 43 | 49 | 55 | 50 | 49 |

**Females**

| | Units (/day) | Obs diet | Modeled diets | | | | | | | | | | | | | |
|---|---|---|---|---|---|---|---|---|---|---|---|---|---|---|---|---|
| | %PP | 34% | 16% | 20% | 25% | 30% | 35% | 40% | 45% | 50% | 55% | 60% | 65% | 70% | 75% | 77% |
| Energy intake | kcal | 2 024 | 2 205 | 2 110 | 2 042 | 1 995 | 1 995 | 1 995 | 1 995 | 1 995 | 1 995 | 2 092 | 2 205 | 2 205 | 2 205 | 2 205 |
| Vitamin A | µg | 608 | 2016 | 751 | 654 | 650 | 650 | 650 | 650 | 650 | 650 | 650 | 650 | 650 | 650 | 650 |
| Vitamin B1 | µg/kcal | 0.56 | 0.63 | 0.67 | 0.71 | 0.67 | 0.65 | 0.64 | 0.63 | 0.63 | 0.63 | 0.64 | 0.66 | 0.72 | 0.66 | 0.70 |
| Vitamin B2 | mg | 1.66 | 3.29 | 3.05 | 3.01 | 2.29 | 2.05 | 1.92 | 1.82 | 1.79 | 1.80 | 1.78 | 1.77 | 1.79 | 1.60 | 1.60 |
| Vitamin B3[3] | mg NE/kcal | 0.016 | 0.023 | 0.020 | 0.021 | 0.019 | 0.017 | 0.016 | 0.014 | 0.013 | 0.012 | 0.012 | 0.012 | 0.013 | 0.014 | 0.014 |
| Vitamin B5 | mg | 5.3 | 9.5 | 9.3 | 9.1 | 7.7 | 7.2 | 6.8 | 6.5 | 6.3 | 6.2 | 6.4 | 7.0 | 7.5 | 6.8 | 7.0 |
| Vitamin B6 | mg | 1.5 | 2.7 | 2.3 | 2.1 | 2.0 | 1.9 | 1.8 | 1.7 | 1.7 | 1.6 | 1.8 | 2.0 | 2.3 | 2.3 | 2.4 |
| Vitamin B9 | µg | 280 | 500 | 460 | 489 | 457 | 448 | 445 | 445 | 448 | 455 | 477 | 507 | 539 | 534 | 556 |
| Vitamin B12 | µg | 4.1 | 18.5 | 9.0 | 8.7 | 5.5 | 5.1 | 5.1 | 5.1 | 5.5 | 5.8 | 5.8 | 5.5 | 4.5 | 5.3 | 4.6 |
| Vitamin C | mg | 84 | 144 | 137 | 127 | 131 | 126 | 126 | 127 | 129 | 131 | 145 | 156 | 164 | 172 | 193 |
| Vitamin D | µg | 2.9 | 5.6 | 6.1 | 6.3 | 5.1 | 4.6 | 4.3 | 3.9 | 3.8 | 3.7 | 3.5 | 3.3 | 2.9 | 2.1 | 1.9 |
| Vitamin E | mg | 11 | 16 | 15 | 14 | 15 | 15 | 15 | 15 | 16 | 16 | 16 | 17 | 14 | 16 | 15 |
| Vitamin K1 | µg | 111 | 231 | 237 | 193 | 213 | 223 | 225 | 226 | 228 | 229 | 238 | 243 | 237 | 251 | 262 |
| Calcium | mg | 892 | 1 552 | 1 711 | 1 505 | 1 239 | 1 123 | 1 066 | 1 037 | 1 054 | 1 068 | 1 069 | 1 044 | 1 025 | 950 | 950 |
| Copper | mg | 1.6 | 4.2 | 2.0 | 2.0 | 1.7 | 1.7 | 1.8 | 1.8 | 1.9 | 1.9 | 2.0 | 2.1 | 2.2 | 2.5 | 2.5 |
| Bioavailable iron[4] | mg | 1.21 | 2.12 | 1.16 | 1.16 | 1.16 | 1.16 | 1.16 | 1.16 | 1.16 | 1.16 | 1.16 | 1.16 | 1.16 | 1.16 | 1.16 |
| *Oral iron* | *mg* | *10* | *16* | *12* | *13* | *11* | *11* | *11* | *11* | *12* | *12* | *12* | *13* | *13* | *15* | *15* |
| *Iron absorption* | *%* | *12%* | *13%* | *9%* | *9%* | *11%* | *11%* | *10%* | *10%* | *10%* | *10%* | *10%* | *9%* | *9%* | *8%* | *8%* |
| Iodine | µg | 145 | 238 | 248 | 238 | 177 | 162 | 157 | 150 | 150 | 150 | 150 | 150 | 150 | 150 | 150 |
| Magnesium | mg | 330 | 471 | 498 | 502 | 401 | 393 | 393 | 393 | 396 | 398 | 415 | 429 | 442 | 522 | 529 |
| Manganese | mg | 3.1 | 3.3 | 3.7 | 4.1 | 3.5 | 3.7 | 3.8 | 3.9 | 4.1 | 4.2 | 4.5 | 4.9 | 5.5 | 5.6 | 5.7 |
| Phosphorus | mg | 1 128 | 1 909 | 1 878 | 1 839 | 1 680 | 1 552 | 1 463 | 1 402 | 1 356 | 1 323 | 1 347 | 1 393 | 1 452 | 1 194 | 1 184 |
| Potassium | mg | 2 906 | 5 380 | 4 974 | 4 536 | 3 905 | 3 789 | 3 747 | 3 711 | 3 710 | 3 725 | 3 935 | 4 207 | 4 252 | 4 537 | 4 656 |
| Selenium | µg | 120 | 166 | 171 | 154 | 133 | 130 | 128 | 122 | 120 | 114 | 119 | 121 | 125 | 171 | 181 |
| Sodium | mg | 3 100 | 2 300 | 2 297 | 2 300 | 2 300 | 2 300 | 2 300 | 2 300 | 2 300 | 2 300 | 2 300 | 2 300 | 2 300 | 2 300 | 2 300 |
| Bioavailable zinc[4] | mg | 3.15 | 4.39 | 4.14 | 3.93 | 3.41 | 3.13 | 2.97 | 2.85 | 2.80 | 2.75 | 2.69 | 2.59 | 2.46 | 2.40 | 2.28 |
| *Oral zinc* | *mg* | *8.8* | *14.6* | *12.6* | *12.4* | *9.9* | *9.2* | *8.9* | *8.8* | *8.8* | *8.8* | *9.0* | *9.2* | *9.0* | *9.4* | *8.9* |
| *Zinc absorption* | *%* | *36%* | *30%* | *33%* | *32%* | *34%* | *34%* | *33%* | *33%* | *32%* | *31%* | *30%* | *28%* | *27%* | *26%* | *26%* |
| Water | g | 2 422 | 2 914 | 3 532 | 2 909 | 2 250 | 2 219 | 2 235 | 2 182 | 2 213 | 2 151 | 2 163 | 2 041 | 2 000 | 3 861 | 4 154 |
| Saturated fatty acids | % EI | 15% | 12% | 12% | 12% | 12% | 12% | 12% | 12% | 12% | 12% | 11% | 10% | 8% | 8% | 7% |
| Atherogenic fatty acids | % EI | 9% | 8% | 8% | 8% | 8% | 7% | 7% | 7% | 7% | 7% | 7% | 6% | 5% | 5% | 4% |
| Linoleic acid | % EI | 3% | 4% | 4% | 5% | 5% | 5% | 5% | 5% | 5% | 5% | 5% | 5% | 4% | 5% | 5% |
| α-linolenic acid | % EI | 0.0% | 1.0% | 1.0% | 1.0% | 1.0% | 1.0% | 1.0% | 1.0% | 1.0% | 1.0% | 1.0% | 1.0% | 1.0% | 1.0% | 1.0% |
| Linoleic / α-linolenic | - | 7 | 4 | 4 | 5 | 5 | 5 | 5 | 5 | 5 | 5 | 5 | 5 | 4 | 5 | 5 |
| EPA+DHA | g | 0.22 | 0.67 | 0.67 | 0.66 | 0.50 | 0.50 | 0.50 | 0.50 | 0.50 | 0.50 | 0.50 | 0.50 | 0.50 | 0.50 | 0.50 |
| Sugar (excluding lactose) | g | 80 | 100 | 100 | 81 | 79 | 79 | 80 | 82 | 85 | 88 | 85 | 75 | 74 | 83 | 96 |
| Protein[5] | g | 76 | 122 | 108 | 110 | 99 | 91 | 83 | 77 | 71 | 66 | 66 | 68 | 72 | 64 | 63 |
| Fiber | g | 19 | 30 | 30 | 30 | 31 | 32 | 33 | 34 | 34 | 35 | 37 | 40 | 44 | 44 | 44 |

[1]Observed data from the third Individual and National Study on Food Consumption French Survey (INCA3), n=1125 (564 males, 561 females).
[2]In the observed diets, the energy and nutrient intakes on a black background are those being outside the range of reference values. In the modeled diets, the intakes on a grey background are those being just equal to (one of) their reference values (i.e., equal to the lower or upper bounds of the nutritional constraints). No reference value or nutritional constraint was considered for vitamin D, as its reference value (15 µg/d) is known to be much too high to be reached through food intake alone (23, 26).
[3]1 mg niacin equivalent (NE) is equal to 1 mg niacin or 60 mg tryptophan.
[4]For bioavailable iron and zinc, lower bounds were not based on current reference values but on lower threshold values ensuring ≤5% deficiency prevalences.
[5]To account for the slightly lower average digestibility of plant protein, protein intake from plants was reduced by 5% when calculating total protein intake.
Atherogenic fatty acids, lauric and myristic and palmitic acids; DHA, docosahexaenoic acid; EI, energy intake; EPA, eicosapentaenoic acid.

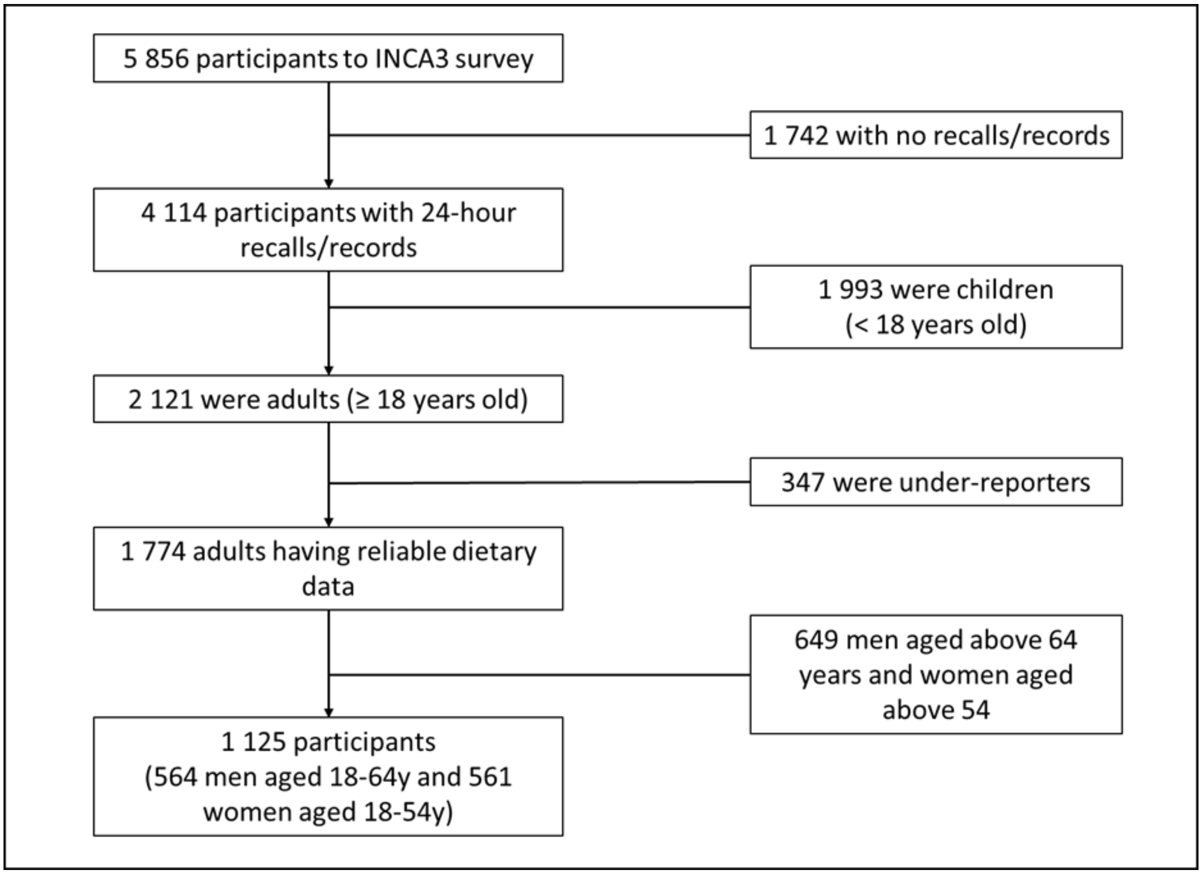

**Fig. S1.** Flow chart explaining the sampling of French participants from the third Individual and National Study on Food Consumption Survey (INCA3) for the present study.

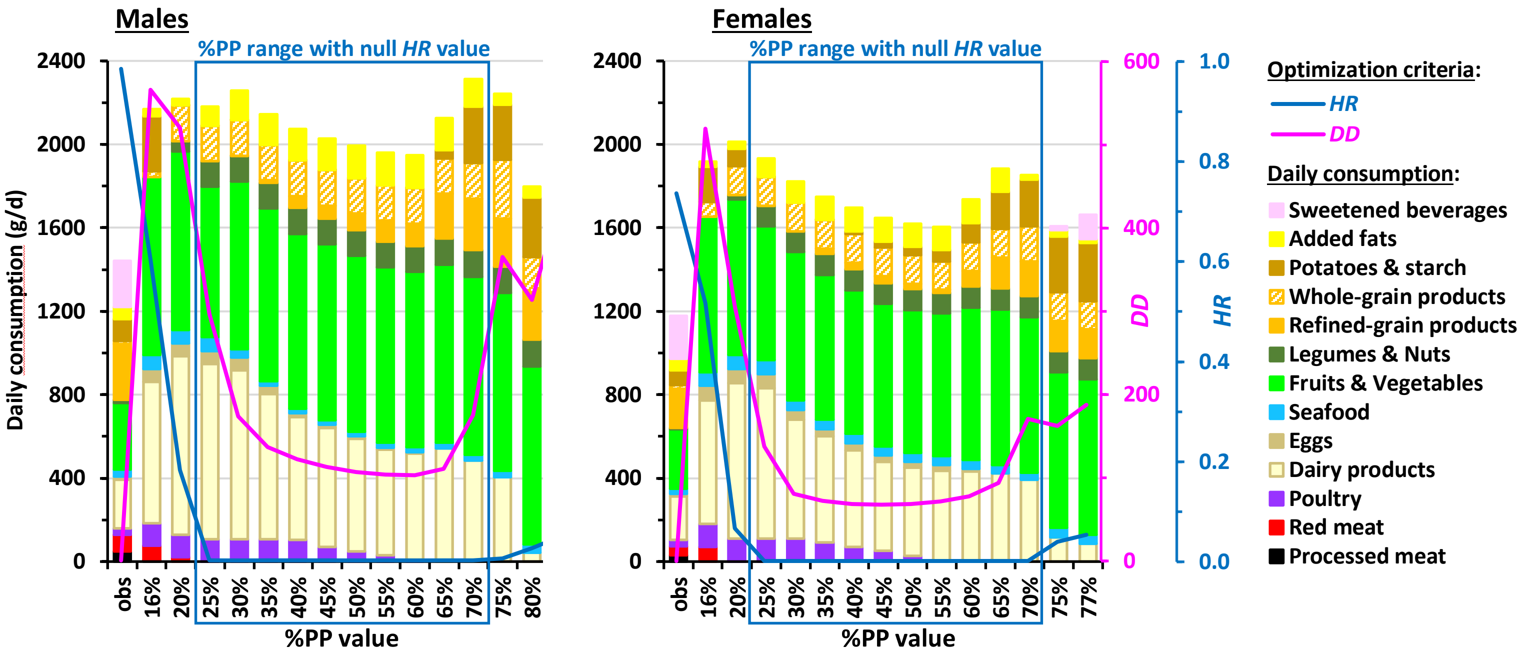

**Fig. S2.** Daily food category consumption in the observed diets (obs) and modeled diets obtained by long-term health risk (*HR*) and diet departure (*DD*) minimization under imposed percentage of plant protein in the diet (%PP) in French males and females. Results are reported for all the adequate %PP values ensuring nutrient adequacy (16%-82% in males and 16%-77% in females), which includes those also ensuring a null *HR* value (25%-70% in both sexes). The bar charts represent the cumulative consumptions of food categories (black axis on the left), and the curves represent the *HR* and *DD* values (blue and pink axes on the right, respectively). For clarity, the 45 modeled food groups are not represented here but grouped into broader categories that are included in *HR* (such as red and processed meats) or represent other protein sources (such as poultry and seafood). Consumptions of water, hot beverages, alcohol and miscellaneous foods are not shown, for clarity. Details about food grouping and consumptions of food categories not shown here are given in Supplemental Tables 1 and 6, respectively.

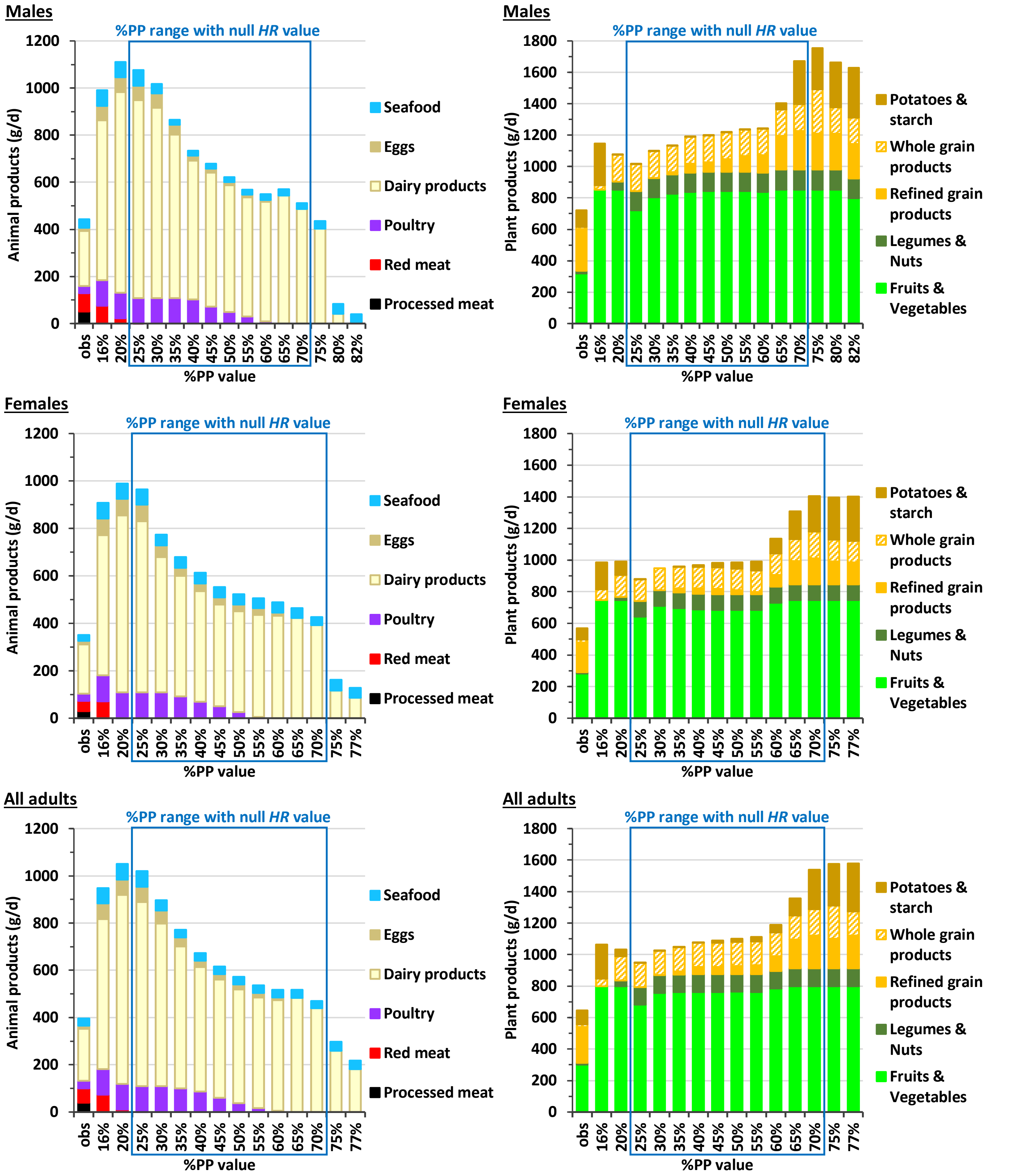

**Fig. S3.** Daily food category consumption in the observed diets (obs) and modeled diets obtained by long-term health risk (*HR*) and diet departure minimization under imposed percentage of plant protein in the diet (%PP) in French males, females and all adults. Results are reported for all the adequate %PP values ensuring nutrient adequacy and model convergence, of which only the 25%-70% range ensured a null *HR* value. The bar charts represent cumulative consumption of the main animal-based or plant-based food categories. Details about food grouping and consumptions of food categories not shown here are given in Supplemental Tables 1 and 6, respectively.

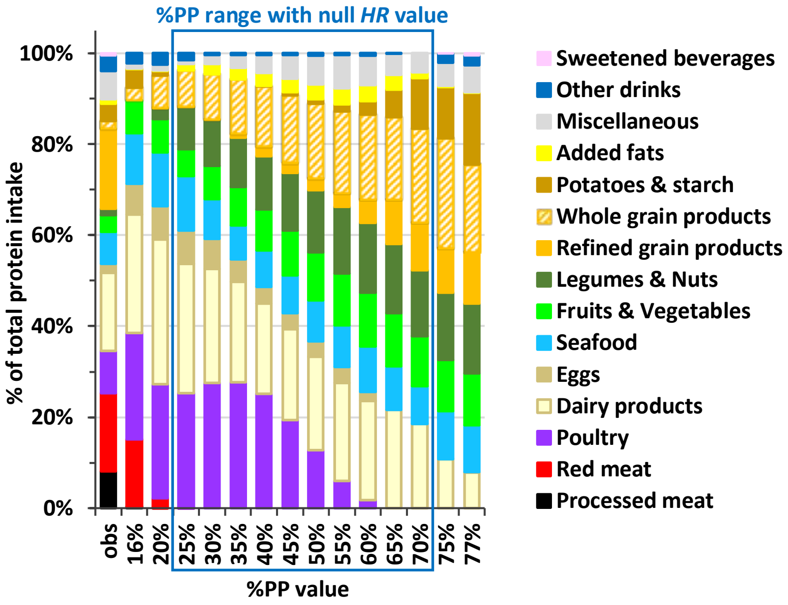

**Fig. S4.** Relative contribution of food categories to protein intake (in % of total protein intake) in the observed diets (obs) and modeled diets obtained by long-term health risk (*HR*) and diet departure minimization under imposed percentage of plant protein in the diet (%PP) in French adults. Results are reported for all the adequate %PP values ensuring nutrient adequacy and model convergence, of which only the 25%-70% range ensured a null *HR* value. See Supplemental Table 1 for the detailed composition of food categories.

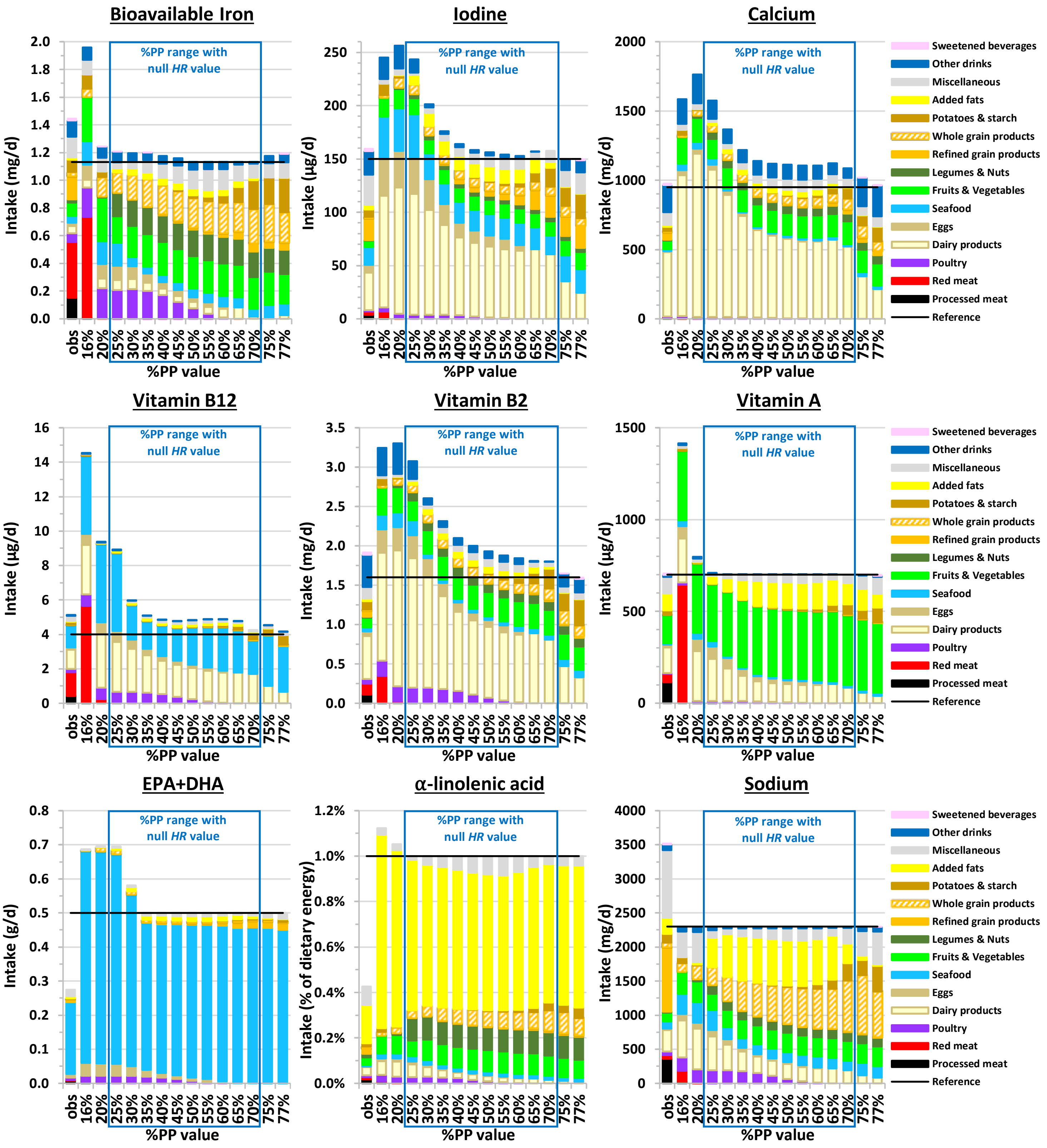

**Fig. S5.** Contribution of food categories to intakes of nutrients that are limiting for a high plant protein percentage in the diet (%PP ≥ 75%) in observed (obs) and modeled diets obtained by long-term health risk (*HR*) and diet departure minimization under imposed %PP value in French adults. Results are reported for all the adequate %PP values ensuring nutrient adequacy and model convergence, of which only the 25%-70% range ensured a null *HR* value. The line represents the constraint for the selected nutrient (lower bound, except for sodium that is upper bound). For bioavailable iron and vitamin A, constraints are represented as the mean between the constraint for males and females. For other nutrients, constraints are similar between sexes. See Supplemental Table 3 for the detailed values of the constraints, and Supplemental Table 1 for the detailed composition of food categories.

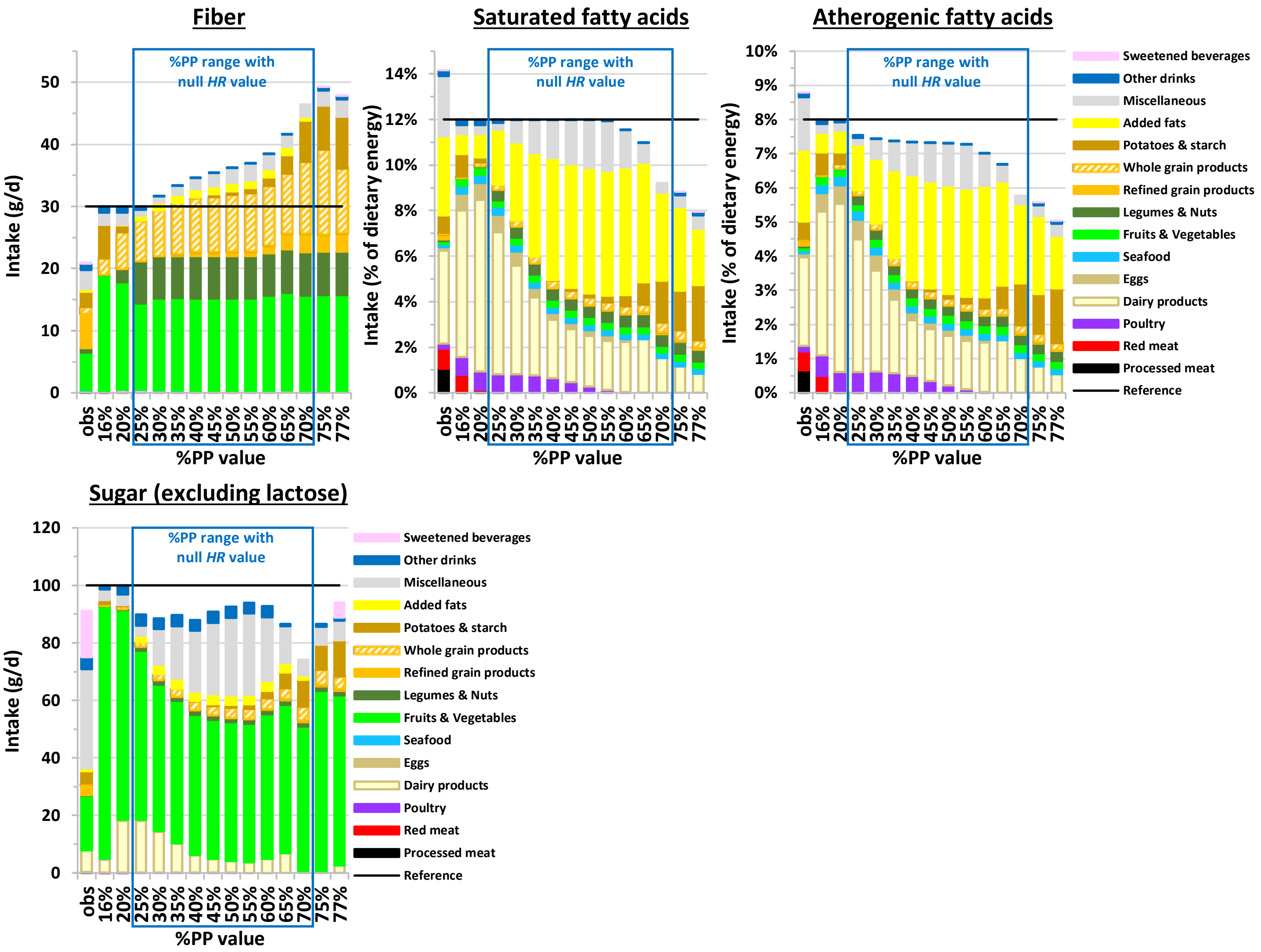

**Fig. S6.** Contribution of food categories to the intakes of nutrients that are limiting for a low plant protein percentage in the diet (%PP ≤ 20%) in observed (obs) and modeled diets obtained by long-term health risk (*HR*) and diet departure minimization under imposed %PP value in French adults. Results are reported for all the adequate %PP values ensuring nutrient adequacy and model convergence, of which only the 25%-70% range ensured a null *HR* value. The line represents the constraint for the selected nutrient (upper bound, except for fiber that is lower bound). See Supplemental Table 1 for the detailed composition of food categories. Atherogenic fatty acids, lauric and myristic and palmitic acids.

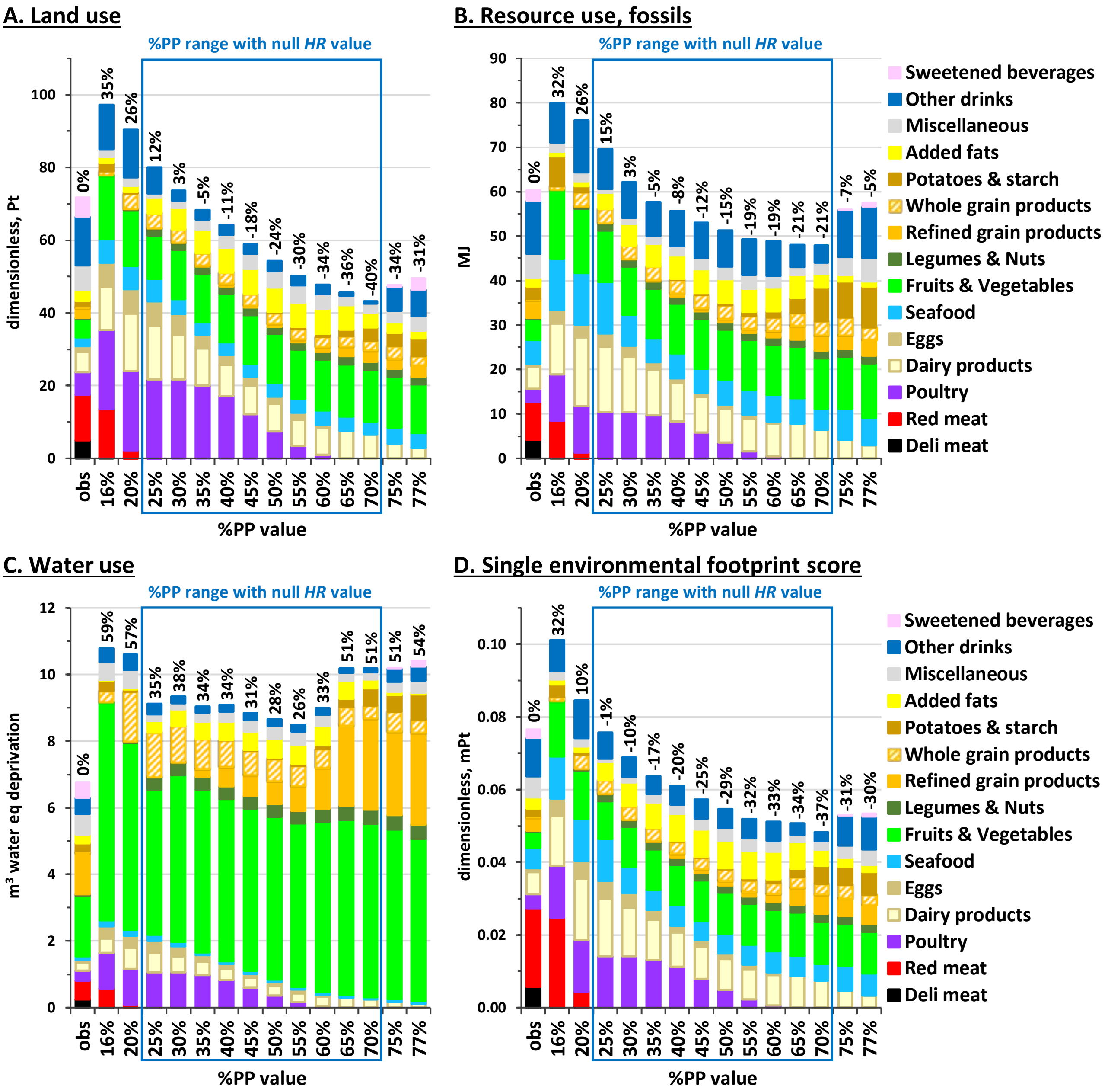

**Fig. S7.** Land use (A), fossils resource use (B), water use (C) and single environmental footprint score aggregating 16 indicators (D) associated with the observed (obs) and modeled diets obtained by long-term health risk (*HR*) and diet departure minimization under imposed percentage of plant protein in the diet (%PP) in French adults. Results are reported for all the adequate %PP values ensuring nutrient adequacy and model convergence, of which only the 25%-70% range ensured a null *HR* value. Sections inside the bars represent the contributions of food categories to the considered environmental indicator, and values above the bars represent the percentage deviation in the environmental indicator from its observed value. See Supplemental Table 1 for the detailed composition of food categories.
